# The development of childhood internalising problems: A meta-analysis of epigenome-wide-association-studies

**DOI:** 10.1101/2025.06.06.25329102

**Authors:** Laura Schellhas, Mannan Luo, Janine F Felix, Christian M Page, Mona Bekkhus, Marcus R Munafò, Luisa Zuccolo, Alexandra Havdahl, Charlotte AM Cecil, Gemma C Sharp

## Abstract

**Background:** Pre- and postpartum environments and genetic effects influence childhood internalising problems, which increase depression risk. DNA methylation (DNAm) may capture some of these effects. We therefore investigated associations between child blood DNAm and internalising problems.

**Method:** We meta-analysed probe and region-level epigenome-wide association studies using data from 3 European birth cohorts (ALSPAC, MoBa, Generation R; maximum n = 3,011) from the Pregnancy And Childhood Epigenetics (PACE) Consortium. DNAm was assessed at birth (cord blood) and age 6 years (peripheral blood). Internalising problems (ages 3 and 6) were reported by mothers using the Child Behaviour Checklist or Strengths and Difficulties Questionnaire. Models were adjusted for age at DNAm assessment, 20 surrogate variables, estimated cell proportions, maternal education, age, smoking, and, in secondary analysis maternal anxiety and depression. Public databases were searched for mental health associations with top CpG sites and regions.

**Results:** No significant probe-level associations were found between cord or peripheral blood DNAm and internalising problems. In region-level analyses, two differentially methylated regions (DMRs) in cord blood were associated with internalising problems at age 3 (annotated to *STK32C*, *MIR886*), and one at age 6 (*PFKFB2*). At age 6, peripheral blood analyses identified two DMRs (*C10orf26* (*WBP1L*), *FAM125A*). Several genes showed prior associations with psychiatric phenotypes, including depression.

**Conclusion:** The higher-powered regional-level analyses revealed more associations than probe-level. Amongst others, we identified a region annotated to *STK32C* that has previously been linked to adolescent depression. Larger samples and refined phenotyping are needed to clarify the role of DNAm in internalising problems.

**KEY POINTS:** 

**What is known?:**

- Childhood internalising problems are influenced by genetic, prenatal, and postnatal environments and are risk factors for later depression.
- DNA methylation (DNAm) has been proposed as a potential biological mechanism linking early exposures to mental health outcomes, though evidence remains inconsistent.

**What is new?:**

- This meta-analysis examined associations between child blood DNAm and internalising problems at multiple time points using both probe- and region-level approaches.
- Region-level analyses, rather than individual CpG sites, revealed associations with internalising problems, identifying regions annotated to genes previously linked to psychiatric phenotypes.

**What is significant for clinical practice?:**

- While findings are preliminary, they suggest that DNAm signatures in early life may help identify biological pathways relevant to childhood internalising problems.
- Larger, longitudinal studies with detailed phenotyping are warranted to confirm these associations before translating DNAm findings into clinical risk markers or interventions.

## INTRODUCTION

Research has shown that childhood internalising problems predict adolescent anxiety and depression, as well as difficulties in academics, social relationships, and overall life satisfaction (Sallis et al., 2019). This underscores the need to understand the early development of internalising problems to prevent the development of more severe problems later in life. In observational studies, intrauterine and early post-natal experiences, such as tobacco use during pregnancy and trauma, emotional or physical abuse in early life, were associated with an increased risk for childhood internalising problems (Duko et al., 2020; LeMoult et al., 2020). Genetic and environmental influences are implicated in the aetiology of internalising disorders, such as anxiety and depression (Kwong et al., 2019). Twin studies suggest that anxiety and depression in childhood are moderately heritable (∼40%) (Polderman et al., 2015) and show a shared genetic liability (Eley, 1999; Gottschalk & Domschke, 2017). Recent genome-wide association studies (GWAS) of anxiety and depression report lower heritability estimates than twin studies, with a SNP-based heritability of 26% to 31% for anxiety (Purves et al., 2020), and 9% for depression (Howard et al., 2019).

Epigenetics may represent a molecular pathway mediating genetic and environmental influences on childhood internalising symptoms (Cortessis et al., 2012; Perera & Herbstman, 2011). Epigenetic changes are influenced by genetic and environmental effects and are involved in regulation of gene activity without modifying the DNA sequence itself (Bird, 2007). The most studied epigenetic modification is DNA methylation (DNAm), which is the addition of a methyl group to a cytosine-phosphate-guanine site (CpGs) in the genome (Bird, 2007). This study investigates whether differences in DNAm at birth and childhood are associated with internalising problems at 3 and 6 years.

Despite numerous epigenome-wide association studies (EWAS) investigating DNAm signals related to risk factors for internalising problems, such as childhood maltreatment (Dunn et al., 2019; Juruena et al., 2021; Parade et al., 2021), only a few small studies have inspected DNAm associations with internalising problems in childhood (Barker et al., 2018). Out of these EWAS, one (n = 190) identified associations between childhood depressive symptoms and three CpGs in childhood peripheral blood (median age = 10.2 years), annotated to genes known to be involved in the stress-response and neurological development (*ID3*, *GRIN1,* and *TPPP)* (Weder et al., 2014). Another small EWAS of 18 monozygotic twin-pairs discordant for adolescent depression (mean age = 16.8 years) found higher buccal cell DNAm at cg07080019, annotated to gene *STK32C*, in the depressed group (Dempster et al., 2014). This association was replicated in a small sample of post-mortem brain tissue discordant for depression (n = 14). Only one small EWAS (n = 256) has investigated associations with internalising problems in young children (18 months), which found no difference in DNAm in cord blood between children with low or high internalising problems (Kallak et al., 2022). None of these studies identified the same CpGs or regions as each other. The incongruent results could be explained by small EWAS being statistically underpowered to capture the complexity of the gene-environment interplay of behavioural phenotypes (Mansell et al., 2019; Non, 2021). Large-scale, longitudinal, harmonized and collaborative efforts are needed to account for the time-varying nature of epigenetic and mental health data across development (Cecil et al., 2023). Therefore, the current study meta-analysed results from EWAS conducted in three European pregnancy and birth cohort studies from the Pregnancy And Childhood Epigenetics (PACE) Consortium, which collectively provided a large sample size (maximum n = 3,011) to investigate the association between DNAm and internalising problems in childhood. As early development is a critical period for the risk for mental health problems later in life, with significant neurological foundations being established within the first three years (Cecil et al., 2023), we measured early manifestations of internalising problems at two timepoints in childhood (ages 3 and 6 years). To address the dynamic nature of DNAm across development and to account for effects of the prenatal- and postnatal environment, DNAm signals were measured at birth (prospective analysis using cord blood, pre-symptom manifestation) and in childhood (cross-sectional analysis using peripheral blood at ages 6).

## METHODS

### Participating cohorts

Three independent birth cohorts contributed data on offspring cord blood DNAm and internalising problems for this study: One UK based cohort (the Avon Longitudinal Study of Parents and Children, ALSPAC; N = 739) (Boyd et al., 2013; Fraser et al., 2013); one Dutch cohort (Generation R; N = 793) (Kooijman et al., 2016; Kruithof et al., 2014) and two datasets from a Norwegian cohort (the Norwegian Mother, Father, and Child Cohort, MoBa1, N = 998 and Moba2, N = 481) (Magnus et al., 2016). Recruitment periods differed among cohorts, from the early 1990s (ALSPAC), 2002-2006 (Generation R) and 1999-2008 (MoBa). At the time of analysis, internalising problems at age 6 were only accessible in ALSPAC and Generation R; thus, MoBa data contributed exclusively to the prospective analysis of internalising problems at age 3. Ethical approval was obtained by local ethics committees and informed consent for the use of data was obtained from all participants. We excluded multiple pregnancies (e.g., twins) and siblings so that each mother was only represented once in the datasets. For more details about the individual cohorts please refer to the Supporting Information File S1.

### Measures

#### DNA methylation

DNAm data was sampled using the lllumina Infinium® HumanMethylation450 (486,425 probes). Cohorts assessed methylation data individually, using their own laboratory methods, quality control, and normalisation. Cord blood and peripheral blood DNAm was assessed through normalised beta values. Probes on SNPs, probes that cross-hybridized according to Chen and colleagues (Chen et al., 2013), and probes on sex chromosomes were excluded (Supporting Information File S1).

#### Childhood internalising problems

Generation R and MoBa assessed child internalising problems by maternal report on the internalising subscale of the Child Behaviour checklist (CBCL-1½-5) (Achenbach & Rescorla, 2000). MoBa1 and MoBa2 used a shortend scale (subset of 9-items) of the full internalising scale. In ALSPAC, internalising problems were assessed using maternal report on the emotional symptoms subscale of the Strengths and Difficulties Questionnaire (SDQ) (Goodman, 1997). Internalising problems scores were z-standardised to ensure compatibility of the scales used in the different cohorts. More information on the measures and their comparability can be found in the Supporting Information File S1.

#### Covariates

All models were adjusted for: age at DNAm assessment; 20 surrogate variables; cell proportions estimated using the Houseman method (Houseman et al., 2012) (prospective analyses: cord blood reference panel (Gervin et al., 2016); cross-sectional analysis: peripheral blood reference panel (Reinius et al., 2012)); maternal education, age and smoking during pregnancy. We conducted secondary analyses stratifying by sex, because sex-specific DNAm differences can persist even after restricting analyses to autosomes and removing probes that are cross-reactive with sex chromosomes (Yousefi et al., 2015). In a sensitivity analysis, we additionally adjusted for maternal anxiety/depression during pregnancy, because parental perinatal depression is robustly associated with childhood internalising problems (Ivanova et al., 2022; Low et al., 2022; Rogers et al., 2020), especially when reported by the same informant (Ivanova et al., 2022), and has been associated with child DNAm (Drzymalla et al., 2021; Robakis et al., 2022). To avoid collinearity, this adjustment was included in a separate model. Maternal anxiety/depression symptoms during pregnancy were z-standardised to harmonise scales across cohorts (Supporting Information File S1).

### Statistical analyses

#### Cohort-specific statistical analyses

Outliers were removed from the methylation data using the Tukey method (values < (25^th^ percentile − 3*interquartile range) and values > (75^th^ percentile + 3*interquartile range)) (Tukey, 1981). For the probe-level analysis, we ran a linear regression at each CpG with offspring DNAm as the exposure and internalising problems in childhood as the outcome using the R-package Limma (Ritchie et al., 2015). Probes were annotated to the human reference genome version 19, build 37 using the annotation data available from the R-package meffil (Suderman et al., 2019). We also conducted a regional analysis, which has higher statistical power than probe-level analyses, in each cohort using the dmrff R-package (Suderman et al., 2018).

#### Meta-analyses

Overall, three different EWAS meta-analysis were run using a probe-level and a regional analysis approach:

1. Prospective analysis age 3: Internalising problems age 3 ∼ Cord blood DNAm (cohorts: ALSPAC, MoBa, Generation R; N = 3,011)
2. Prospective analysis age 6: Internalising problems age 6 ∼ Cord blood DNAm (cohorts: ALSPAC, Generation R; N = 1,601)
3. Cross-sectional analysis age 6: Internalising problems age 6 ∼ Peripheral DNAm age 6 (cohorts: ALSPAC, Generation R; N = 1,121)

For each of these analyses, we ran one main model, two models stratified by sex, and one sensitivity model additionally adjusted for maternal anxiety and depression.

##### Probe-level analysis

Quality checks on the cohort and meta-analysis results were conducted (QQ- and correlation plots) (Sharp et al., 2021; Van der Most et al., 2017). Results of the probe-level analysis were meta-analysed with fixed effect estimates, weighted by the inverse of the variance using METAL (Willer et al., 2010). To account for multiple testing, *p*-values were adjusted using a 5% false discovery rate (FDR) (Benjamini & Hochberg, 1995) and values < 0.05 were considered as evidence for statistical significance. A leave-one-cohort-out analysis was performed on the CpGs with the strongest evidence for a statistical association (as indicated by the smallest *p*-value) using the R package *metafor* (Viechtbauer, 2010). A change in direction of the estimate, moved toward the null by more than 20%, or a confidence interval that included 0 after removal of a single cohort were considered as indication for results being driven by a single cohort. A shadow meta-analysis was conducted independently by researchers at Generation R.

##### Differentially methylated regions (DMR) analysis

The meta-analysis of the DMR cohort results was conducted using the *dmrff.meta* function within the dmrff R-package, which uses an inverse-variance weighted fixed effects approach (Suderman et al., 2018). A region was defined as having at least two CpGs that are less than 500 bp apart, show the same direction of effect and a Bonferroni adjusted *p*-value (*P*_Bonferroni_) < 0.05 (Shaffer, 1995).

##### Database look-up

To contextualise findings from the probe and DMR analyses, we consulted publicly available databases: GeneCards (https://www.genecards.org/), NHGRI-EBI GWAS catalogue (GWAS catalogue; https://www.ebi.ac.uk/gwas/) and EWAS catalog (http://www.ewascatalog.org/) and manually screened for mental health phenotypes. We defined mental health phenotypes as psychiatric diseases or symptoms of psychiatric diseases (excluding substance abuse). Childhood abuse was also included, given its frequent investigation in relation to DNAm (Dunn et al., 2019; Juruena et al., 2021; Mehta et al., 2023; Parade et al., 2021) and its established role as a risk factor for later depression and anxiety (Gardner et al., 2019; LeMoult et al., 2020; McKay et al., 2022).

## RESULTS

### Sample characteristics

Overall, offspring and maternal characteristics were similar across the cohorts (Table 1).

**Table 1.**
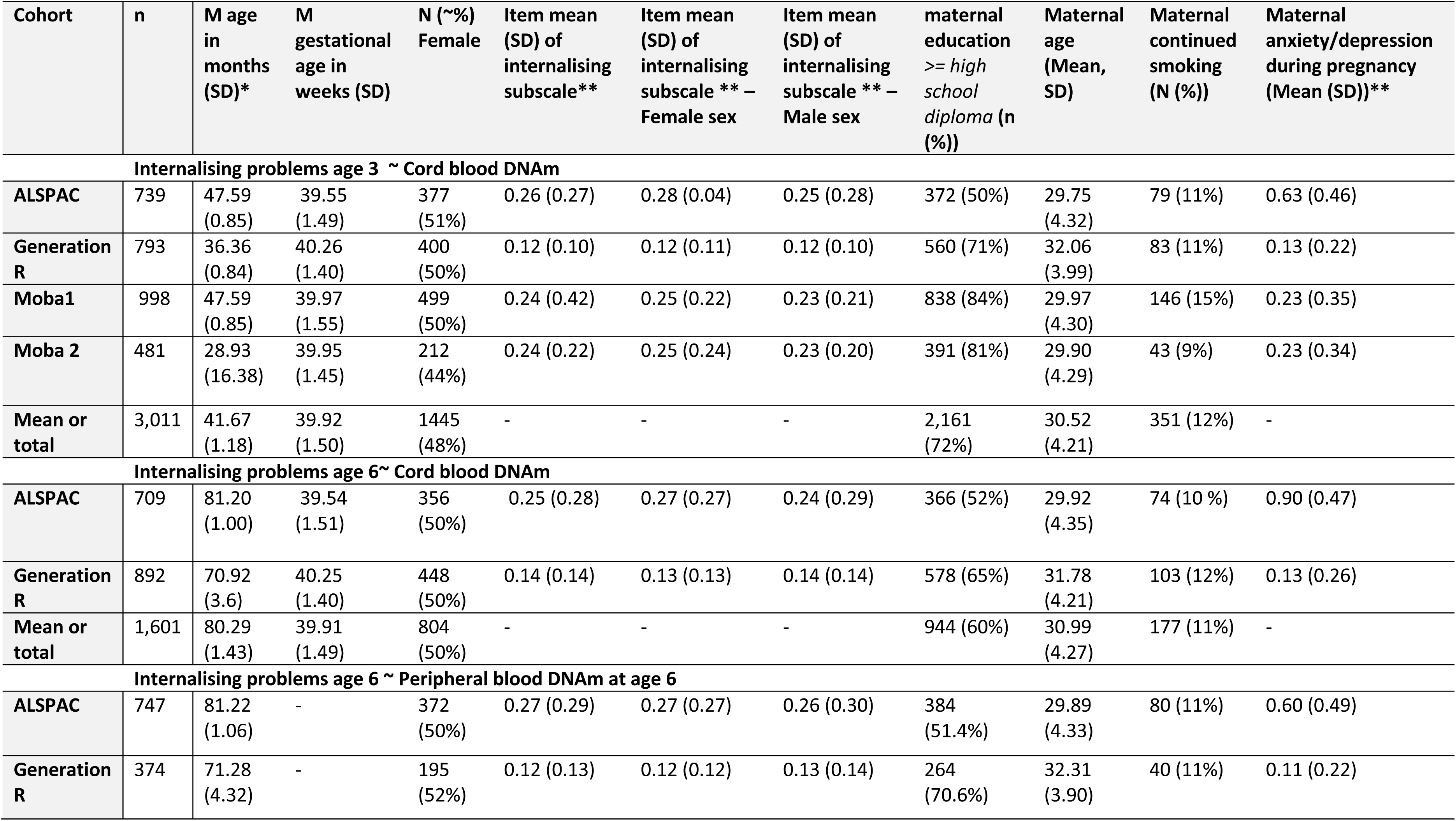
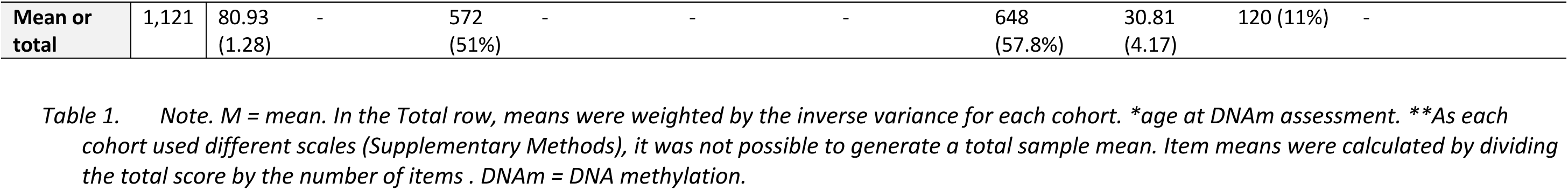
Sample characteristics.

| Cohort | n | M age in months (SD)* | M gestational age in weeks (SD) | N (~%) Female | Item mean (SD) of internalising subscale** | Item mean (SD) of internalising subscale ** – Female sex | Item mean (SD) of internalising subscale ** – Male sex | maternal education >= high school diploma (n (%)) | Maternal age (Mean, SD) | Maternal continued smoking (N (%)) | Maternal anxiety/depression during pregnancy (Mean (SD))** |
| --- | --- | --- | --- | --- | --- | --- | --- | --- | --- | --- | --- |
| **Internalising problems age 3 ~ Cord blood DNAm** |  |  |  |  |  |  |  |  |  |  |  |
| **ALSPAC** | 739 | 47.59 (0.85) | 39.55 (1.49) | 377 (51%) | 0.26 (0.27) | 0.28 (0.04) | 0.25 (0.28) | 372 (50%) | 29.75 (4.32) | 79 (11%) | 0.63 (0.46) |
| **Generation R** | 793 | 36.36 (0.84) | 40.26 (1.40) | 400 (50%) | 0.12 (0.10) | 0.12 (0.11) | 0.12 (0.10) | 560 (71%) | 32.06 (3.99) | 83 (11%) | 0.13 (0.22) |
| **Moba1** | 998 | 47.59 (0.85) | 39.97 (1.55) | 499 (50%) | 0.24 (0.42) | 0.25 (0.22) | 0.23 (0.21) | 838 (84%) | 29.97 (4.30) | 146 (15%) | 0.23 (0.35) |
| **Moba 2** | 481 | 28.93 (16.38) | 39.95 (1.45) | 212 (44%) | 0.24 (0.22) | 0.25 (0.24) | 0.23 (0.20) | 391 (81%) | 29.90 (4.29) | 43 (9%) | 0.23 (0.34) |
| **Mean or total** | 3,011 | 41.67 (1.18) | 39.92 (1.50) | 1445 (48%) | - | - | - | 2,161 (72%) | 30.52 (4.21) | 351 (12%) | - |
| **Internalising problems age 6~ Cord blood DNAm** |  |  |  |  |  |  |  |  |  |  |  |
| **ALSPAC** | 709 | 81.20 (1.00) | 39.54 (1.51) | 356 (50%) | 0.25 (0.28) | 0.27 (0.27) | 0.24 (0.29) | 366 (52%) | 29.92 (4.35) | 74 (10 %) | 0.90 (0.47) |
| **Generation R** | 892 | 70.92 (3.6) | 40.25 (1.40) | 448 (50%) | 0.14 (0.14) | 0.13 (0.13) | 0.14 (0.14) | 578 (65%) | 31.78 (4.21) | 103 (12%) | 0.13 (0.26) |
| **Mean or total** | 1,601 | 80.29 (1.43) | 39.91 (1.49) | 804 (50%) | - | - | - | 944 (60%) | 30.99 (4.27) | 177 (11%) | - |
| **Internalising problems age 6 ~ Peripheral blood DNAm at age 6** |  |  |  |  |  |  |  |  |  |  |  |
| **ALSPAC** | 747 | 81.22 (1.06) | - | 372 (50%) | 0.27 (0.29) | 0.27 (0.27) | 0.26 (0.30) | 384 (51.4%) | 29.89 (4.33) | 80 (11%) | 0.60 (0.49) |
| **Generation R** | 374 | 71.28 (4.32) | - | 195 (52%) | 0.12 (0.13) | 0.12 (0.12) | 0.13 (0.14) | 264 (70.6%) | 32.31 (3.90) | 40 (11%) | 0.11 (0.22) |
| **Mean or total** | 1,121 | 80.93<br>(1.28) | - | 572<br>(51%) | - | - | - | 648<br>(57.8%) | 30.81<br>(4.17) | 120 (11%) | - |

The sample of the prospective meta-analysis at age 3 contained more mothers with at least a high school diploma (69%) than without, and this pattern was consistent across cohorts, except for ALSPAC, where having a high school diploma or not was approximately equally distributed (Table 1). On average, 11% of mothers in the samples smoked in the third trimester or throughout pregnancy (Table 1). Across samples, mothers were on average 31 years of age at the birth of the study child (range: 29.8 to 32.3 years; Table 1). Female and male sex children showed similarly low levels of internalising problems across cohorts (Table 1). Across cohorts and time points there was evidence for a statistically significant positive correlation between maternal anxiety and depressive symptoms during pregnancy and offspring internalising problems, yet the magnitude of associations was rather small (correlation range: 0.13 to 0.23; Table S1). As expected, there was evidence for internalising problems at age 3 being positively associated with internalising problems at the age of 6, but only of moderate size, indicating that internalising problems change from age 3 to 6 (correlation range: 0.39 to 0.56, Table S1).

### Quality control checks

The cohort and meta-analyses quality control checks did not indicate evidence for systematic confounding (Supporting Information File S1; Figure S1 to Figure S29). As expected, correlations between the regression coefficients across models were higher when DNAm was assessed in the same tissue (cord blood *r* = 0.282) than across tissues (cord and peripheral blood *r* = 0.003; Supporting Information File S1, Figure S30).

### Probe-level meta-analyses

#### Prospective probe-level analyses in cord blood

The main analyses revealed no statistically significant association between DNAm in cord-blood and internalising problems at ages 3 or 6 after correcting for multiple testing (Table 2). These results remained consistent after adjusting the main model for maternal anxiety and depression. In the secondary sex-stratified analyses, only the female-specific analyses showed statistical evidence for associations between DNAm at one CpG site in cord blood and internalising problems at age 6 (cg26668632, nearest gene *IFNGR1*; estimate = -8.76, 95%CI: -11.78 to -5.74, *p*-value = 1.42x10^-8^). The direction of effect for cg26668632 was consistent in the female and male sex strata, with a larger magnitude in the female than male subgroup and did survive the leave-one-out analysis (Supporting Information File S1, Figure S23).

**Table 2.** A summary of results of each EWAS model from the probe-level analysis.

| Model | N CpGs with FDR-corrected P-value <0.05 | N CpGs (%) surviving leave-one-out analysis | Meta-analysis sample size | Genomic inflation factor ($\lambda$)** |
| --- | --- | --- | --- | --- |
| **Prospective analysis: Internalising problems age 3 ~ Cord blood DNAm (N = 3,011)** |  |  |  |  |
| All offspring (minimally adjusted)* | 0 | n.a. | 3011 | 1.01 |
| All offspring (adjusted for covariates) | 0 | n.a. | 3011 | 1.00 |
| Female sex offspring (adjusted for covariates) | 0 | n.a. | 1452 | 1.02 |
| Male sex offspring (adjusted for covariates) | 0 | n.a. | 1559 | 0.99 |
| All offspring (adjusted for covariates and maternal anx/dep) | 0 | n.a. | 2987 | 1.00 |
| **Prospective analysis: Internalising problems age 6 ~ Cord blood DNAm (N = 1,601)** |  |  |  |  |
| All offspring (minimally adjusted)* | 0 | n.a. | 1601 | 1.02 |
| All offspring (adjusted for covariates) | 0 | n.a. | 1601 | 1.03 |
| Female sex offspring (adjusted for covariates) | 1 | 1 (100%) | 804 | 1.02 |
| Male sex offspring (adjusted for covariates) | 0 | n.a. | 797 | 1.06 |
| All offspring (adjusted for covariates and maternal anx/dep) | 0 | n.a. | 1601 | 1.03 |
| **Cross-sectional analysis: Internalising problems age 6 ~ Childhood DNAm age 6 (N = 1,121)** |  |  |  |  |
| All offspring (minimally adjusted)* | 0 | n.a. | 1601 | 0.99 |
| All offspring (adjusted for covariates) | 0 | n.a. | 1136 | 0.98 |
| Female sex offspring (adjusted for covariates) | 2 | 1 (50%) | 572 | 1.02 |
| Male sex offspring (adjusted for covariates) | 0 | n.a. | 564 | 0.94 |
| All offspring (adjusted for covariates and maternal anx/dep) |  | n.a. | 1136 | 0.98 |

Note. \*only adjusted for estimated cell counts and 20 surrogate variables. Covariates: maternal age, maternal smoking, maternal education, offspring age, estimated cell counts and 20 surrogate variables. \*\* The genomic inflation factor ( $\lambda$ ) estimates the extent of bulk inflation of EWAS p-values and the excess false positive rate. 1 = no inflation; > 1 some evidence of inflation.

#### Cross-sectional probe-level analysis in peripheral blood

No association between DNAm in peripheral blood age 6 and internalising problems at the age of 6 were found in the main model and the model additionally adjusted for maternal anxiety and depression (Table 2). Of the sex-stratified analyses, only the female-specific model found DNAm at two CpGs in peripheral blood in childhood (cg07283896; estimate = 6.10, 95%CI: 4.20 to 8.00; *p*-value = 3.20 x 10^-10^; cg08884410; estimate = 8.59, 95%CI: 5.41 to 11.77, *p*-value = 1.11 x 10^-7^; no annotated genes; Table S3) to be associated with internalising problems at age 6. Of those internalising problems-associated CpGs, only cg08884410 survived the leave-one-out analysis (Supporting Information File S1, Figure S24 and S25). The association with internalising problems at cg08884410 showed a consistent direction of effect across time points and cohorts in peripheral blood of female-sex children (Table S3). Only cg07283896 showed a different direction of effect across the sex strata and time points (Table S3). The female subgroup had slightly more children (+8), while the male subgroup showed greater variability in effect sizes across studies (*I²* = 73.9% for males; *I²* = 0% for females).

### Differentially methylated regions meta-analysis

An overview of the DMR meta-analyses results can be found in Table 3.

**Table 3.**
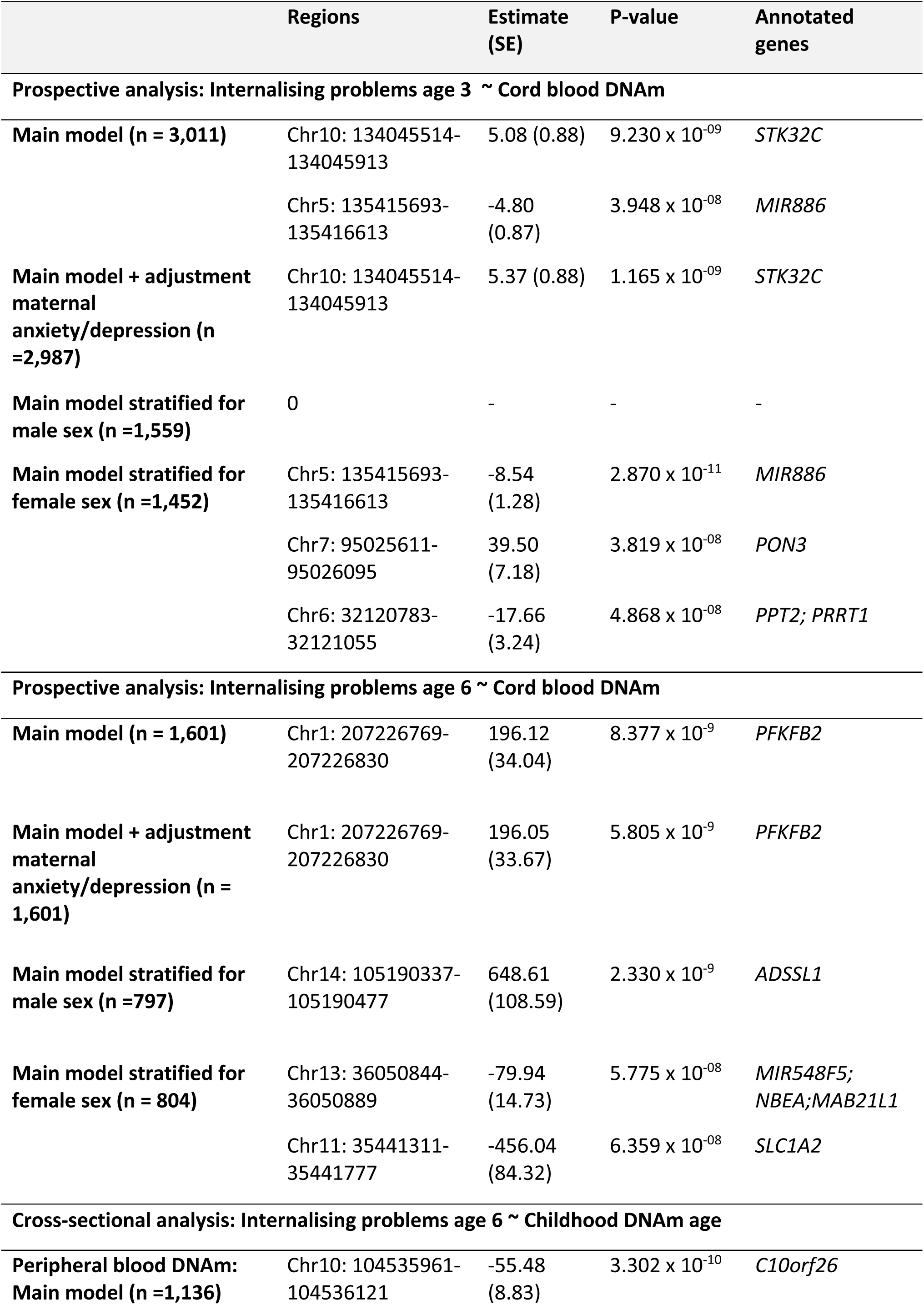
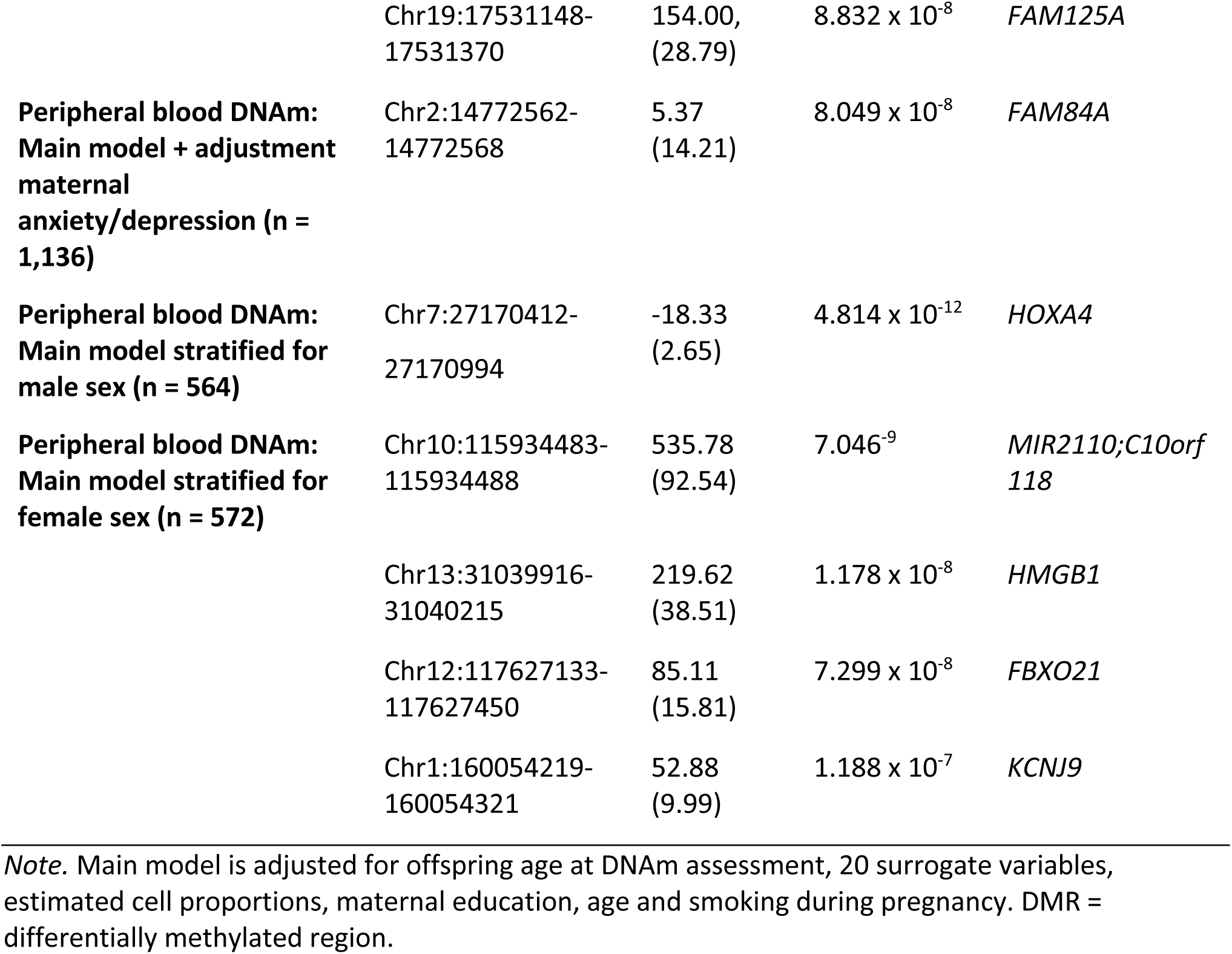
Result overview of the DMR meta-analyses.

|  | Regions | Estimate (SE) | P-value | Annotated genes |
| --- | --- | --- | --- | --- |
| **Prospective analysis: Internalising problems age 3 ~ Cord blood DNAm** |  |  |  |  |
| **Main model (n = 3,011)** | Chr10: 134045514-134045913 | 5.08 (0.88) | $9.230 \times 10^{-09}$ | *STK32C* |
| | Chr5: 135415693-135416613 | -4.80 (0.87) | $3.948 \times 10^{-08}$ | *MIR886* |
| **Main model + adjustment maternal anxiety/depression (n = 2,987)** | Chr10: 134045514-134045913 | 5.37 (0.88) | $1.165 \times 10^{-09}$ | *STK32C* |
| **Main model stratified for male sex (n = 1,559)** | 0 | - | - | - |
| **Main model stratified for female sex (n = 1,452)** | Chr5: 135415693-135416613 | -8.54 (1.28) | $2.870 \times 10^{-11}$ | *MIR886* |
| | Chr7: 95025611-95026095 | 39.50 (7.18) | $3.819 \times 10^{-08}$ | *PON3* |
| | Chr6: 32120783-32121055 | -17.66 (3.24) | $4.868 \times 10^{-08}$ | *PPT2; PRRT1* |
| **Prospective analysis: Internalising problems age 6 ~ Cord blood DNAm** |  |  |  |  |
| **Main model (n = 1,601)** | Chr1: 207226769-207226830 | 196.12 (34.04) | $8.377 \times 10^{-9}$ | *PFKFB2* |
| **Main model + adjustment maternal anxiety/depression (n = 1,601)** | Chr1: 207226769-207226830 | 196.05 (33.67) | $5.805 \times 10^{-9}$ | *PFKFB2* |
| **Main model stratified for male sex (n = 797)** | Chr14: 105190337-105190477 | 648.61 (108.59) | $2.330 \times 10^{-9}$ | *ADSSL1* |
| **Main model stratified for female sex (n = 804)** | Chr13: 36050844-36050889 | -79.94 (14.73) | $5.775 \times 10^{-08}$ | *MIR548F5; NBEA; MAB21L1* |
| | Chr11: 35441311-35441777 | -456.04 (84.32) | $6.359 \times 10^{-08}$ | *SLC1A2* |
| **Cross-sectional analysis: Internalising problems age 6 ~ Childhood DNAm age** |  |  |  |  |
| **Peripheral blood DNAm: Main model (n = 1,136)** | Chr10: 104535961-104536121 | -55.48 (8.83) | $3.302 \times 10^{-10}$ | *C10orf26* |
|  | Chr19:17531148-17531370 | 154.00,<br>(28.79) | 8.832 x 10<sup>-8</sup> | *FAM125A* |
| **Peripheral blood DNAm:**<br>**Main model + adjustment**<br>**maternal**<br>**anxiety/depression (n =**<br>**1,136)** | Chr2:14772562-14772568 | 5.37<br>(14.21) | 8.049 x 10<sup>-8</sup> | *FAM84A* |
| **Peripheral blood DNAm:**<br>**Main model stratified for**<br>**male sex (n = 564)** | Chr7:27170412-27170994 | -18.33<br>(2.65) | 4.814 x 10<sup>-12</sup> | *HOXA4* |
| **Peripheral blood DNAm:**<br>**Main model stratified for**<br>**female sex (n = 572)** | Chr10:115934483-115934488 | 535.78<br>(92.54) | 7.046<sup>-9</sup> | *MIR2110;C10orf118* |
|  | Chr13:31039916-31040215 | 219.62<br>(38.51) | 1.178 x 10<sup>-8</sup> | *HMGB1* |
|  | Chr12:117627133-117627450 | 85.11<br>(15.81) | 7.299 x 10<sup>-8</sup> | *FBXO21* |
|  | Chr1:160054219-160054321 | 52.88<br>(9.99) | 1.188 x 10<sup>-7</sup> | *KCNJ9* |

*Note.* Main model is adjusted for offspring age at DNAm assessment, 20 surrogate variables, estimated cell proportions, maternal education, age and smoking during pregnancy. DMR = differentially methylated region.

#### Prospective DMR analyses in cord-blood

At the age of 3, the main model showed evidence for DMRs in cord-blood, annotated to genes *STK32C* and *MIR886,* being associated with internalising problems. Only the association with the *STK32C* annotated region remained after adjustment for maternal anxiety and depressive symptoms during pregnancy (Table 3). In the sex-stratified models, no regions were identified in the male stratum, but three regions were identified in the models restricted to females. One of these regions (at *MIR886* (*VTRNA2-1*)) was the same as the region identified in the unstratified analyses using the complete group. The other two regions are annotated to the genes *PON3* and *PPT2; PRRT1* (Table 3). At the age of 6, one region in cord blood, that is annotated to gene *PFKFB2,* was associated with internalising problems (Table 3). This region was also differentially methylated in the model additionally adjusted for maternal anxiety and depression symptoms during pregnancy. Each of the sex-strata showed associations with two different DMRs in cord blood. In females, the regions are annotated to genes *MIR548F5; NBEA; MAB21L1* and *SLC1A2*. In males, only one of the regions was annotated to a gene (*ADSSL1*; Table 3). None of the internalising problems associated DMRs in cord blood were overlapping between ages 3 and 6 years (Table 3).

#### Cross-sectional DMR analysis in peripheral blood

In the cross-sectional analysis, the main model showed evidence for two DMRs in peripheral blood, mapped to genes *C10orf26* and *FAM125A*, being associated with internalising problems at age 6. The model adjusted for maternal anxiety and depression indicated an internalising problem associated DMR annotated to gene *FAM84A* (Table 3). The sex-stratified analyses showed evidence for 5 DMRs in the female (annotated genes *MIR2110 (C10orf118)*, *HMGB1*, *FBXO21* and *KCNJ9)*, and 1 DMR in the male models (annotated gene *HOXA4)*, to be associated with internalising problems at age 6.

### Database look-up

A summary of the mental health phenotypes that appeared in the database search of the differentially methylated CpG sites and regions, and their corresponding annotated genes, can be found in Table 4. For an overview of all associated phenotypes please refer to Supporting Information Files S2 (GeneCards), S3 (GWAS catalogue) and S4 (EWAS catalog).

**Table 4.** Result overview of the database look-up.

| Gene | Mental Health Phenotype | Data Source |
| --- | --- | --- |
| **Prospective analysis: Internalising problems age 3 ~ Cord blood DNAm (n = 3,011)** |  |  |
| *STK32C* | Adolescent depression (DNAm studies), risk-taking behavior, ADHD, trauma exposure | GeneCards, GWAS, EWAS |
| *MIR886 (VTRNA2-1)* | Response to antidepressants, bipolar disorder, schizophrenia, borderline personality disorder, ADHD | GWAS Catalogue |
| *PON3, PPT2, PRRT1* | No links with mental health phenotypes | GeneCards, GWAS catalogue, EWAS catalog |
| **Prospective analysis: Internalising problems age 6 ~ Cord blood DNAm (n = 1,601)** |  |  |
| *cg26668632* | No links with mental health phenotypes | EWAS Catalog |
| *IFNGR1* | Childhood abuse | EWAS Catalog |
| *PFKFB2* | Schizophrenia, externalizing behaviors | GWAS Catalogue |
| *SLC1A2* | Schizophrenia | GWAS Catalogue |
| *NBEA* | Antipsychotic treatment response in schizophrenia, post-traumatic stress disorder, externalizing behavior | GWAS Catalogue |
| *ADSSL1* | Schizophrenia, childhood abuse | EWAS Catalog |
| *MIR548F5, MAB21L1, SLC1A2* | No links with mental health phenotypes | GeneCards, GWAS catalogue, EWAS catalog |
| **Cross-sectional analysis: Internalising problems age 6 ~ Childhood DNAm age 6 (n = 1,121)** |  |  |
| *C10orf26 (WBP1L)* | Schizophrenia, anxiety disorder, worry, nap duration in severe depression | GWAS Catalogue |
| *KCNJ9* | Depression, schizophrenia | GWAS Catalogue, EWAS Catalog |
| *MIR2110* | Schizophrenia, childhood abuse | EWAS Catalog |
| *FBXO21* | Insomnia | GWAS Catalogue |
| *cg07283896, cg08884410* | No links with mental health phenotypes | EWAS Catalog |
| *FAM84A (LRATD)* | Sex-dependent genetic interactions, schizophrenia, bipolar disorder, major depressive disorder | GWAS Catalogue |
| *FAM125A (MVB12), HOXA4, HMGB1* | No links with mental health phenotypes | GeneCards, GWAS catalogue, EWAS catalog |

#### Prospective analysis in cord-blood

The differentially methylated CpG site (cg26668632, annotated to *IFNGR1*) had no reported mental health associations, though another *IFNGR1* site (cg13370280) was previously linked to childhood abuse (EWAS catalog).

*STK32C*, was previously associated with adolescent depression (GeneCards), and linked to risk-taking, ADHD, and trauma exposure (GWAS catalogue), with additional ADHD-related methylation findings (EWAS catalog). *MIR886* (*VTRNA2-1*), showed GWAS catalogue associations with several psychiatric phenotypes, but no links in GeneCards or the EWAS catalog. *PON3* and *PPT2/PRRT* showed no associations across databases. No mental health associations for *IFNGR1* were found in GeneCards or the GWAS catalogue. *PFKFB2,* was associated with schizophrenia and externalizing behaviours (GWAS catalogue), with no links in the other databases. *SLC1A2* and *NBEA*, were linked to schizophrenia and other psychiatric phenotypes (GWAS catalogue). *ADSSL1,* had no associations in GeneCards or the GWAS catalogue, but EWAS catalog linked DNAm at CpGs annotated to *ADSSL1* with schizophrenia and child abuse. *MIR548F5* and *MAB21L1* showed no associations across databases. A comprehensive summary of these associations is provided in Table 4.

#### Cross-sectional analysis in peripheral blood

No mental health-related associations were reported in the EWAS Catalog for the differentially methylated CpG sites cg07283896 and cg08884410, both of which lacked gene annotations.

*C10orf26* (*WBP1L*), showed links with schizophrenia, anxiety disorder, worry, and nap duration in severe depression in the GWAS catalogue. *FAM84A* (*LRATD*), showed associations with sex-dependent genetic interactions related to schizophrenia, bipolar disorder, and major depressive disorder in the GWAS Catalogue. DNAm at *MIR2110* has been associated with schizophrenia and childhood abuse in the EWAS catalog, though no associations appeared in the GWAS Catalogue or GeneCards. *KCNJ9* was associated with depression (GWAS Catalogue) and schizophrenia (EWAS Catalog). *FBXO21* was linked to insomnia in the GWAS Catalogue. No mental health-related associations were reported for *FAM125A* (*MVB12*), *HOXA4*, or *HMGB1* across any of the examined databases. A detailed overview of these associations is presented in Table 4.

## DISCUSSION

### Summary and interpretation of findings

The results of this study provide some evidence for a link between DNAm and internalising problems in childhood. Although the probe-level analyses did not identify DNAm at single probes to be associated with childhood internalising problems, the higher powered regional-based analyses revealed several internalising problems associated-DMRs in cord- and peripheral blood.

Results differed depending on the timing of assessments, both for DNAm (cord blood at birth vs. peripheral blood in childhood) and for internalising problems (assessed at ages 3 and 6 years). This aligns with a study from the PACE consortium, which found associations specific to the timing of DNAm assessment (birth vs childhood) for most outcomes, such as ADHD and general psychopathology (Neumann et al., 2025). The incongruence could reflect false positives/negatives from sample size differences, or different confounding structures at different developmental stages. Comparing DNAm in cord blood with peripheral blood in childhood may be problematic due to cell composition and gene regulation differences (Martino et al., 2011; Mulder et al., 2020). This is supported by the higher correlation in cord blood DNAm despite different internalising problem assessment times (ages 3 and 6) compared to the analysis of both cord- and peripheral blood at age 6 (Supporting Information File 1). The incongruence in results could also indicate that DNAm at different CpGs/regions might be associated with internalising problems at different ages, as suggested by the moderate correlation of internalising problems at ages 3 and 6 (correlation range: 0.39 to 0.56, Table S1). Also, different mechanisms may come into play after birth, such as parenting behaviours, health practices, or socioeconomic position, which may influence postnatal DNAm. Except for the association between DNAm at *STK32C* and internalising problems at age 3, no congruence in results was observed between the maternal anxiety and depression adjusted and unadjusted models, indicating that maternal anxiety and depression may play a more complex role in shaping DNAm patterns. The association between DNAm at *MIR886*( *VTRNA2-1*) and child internalising problems at age 3 came up only after adjusting for maternal depression. As *MIR886* (*VTRNA2-1*) is 75% maternally imprinted and has previously been linked to perinatal factors, such as maternal age and season of conception (Carpenter et al., 2018), this locus may reflect sensitivity to the broader prenatal environment. However, interpretations should remain cautious, as these associations likely reflect complex interactions between biological, environmental, and social factors rather than direct causal pathways. Further research is needed to understand the functional effects of the DMRs in a developmental context and clarify timing effects (Neumann et al., 2025).

The database search showed that many genes associated with the DMRs identified in this study have known links to mental health phenotypes. *STK32C*, for example, that showed differential DNAm in cord-blood to be associated with internalising problems age 3, showed various links with mental health phenotypes in the databases, including adolescent depression (Dempster et al., 2014). Another study found adolescents at risk for generalized anxiety disorder (GAD; N = 221) to show different DNAm levels at cg16333992 in *STK32B*, a gene from the same Serine/Threonine Kinase 32 (*STK32*) family (Ciuculete et al., 2018). Besides *STK32C*, GeneCards and the GWAS catalogue linked several other genes from the DMR analysis to depression (*MIR886* (*VTRNA2*-*1*), *KCNJ9*, *FAM84A* (*LRATD1*); Table 4) and anxiety disorder (*C10orf26* (*WBP1L*)).

Sex-stratified analyses revealed more DNAm associations with internalising problems in females, despite no observed sex differences in internalising problems in our study (Table 1). These findings should be interpreted with caution, as the direction of the probe-level effects was the same for both sexes, and the female group was slightly larger with lower variability (Supporting Information File S1, Table S3).

Prior research highlights sex differences in autosomal DNAm and suggests sex-specific epigenetic signatures could inform precision psychiatry (Grant et al., 2022; Mulder et al., 2021; Müller et al., 2021; Tesfaye et al., 2024; Xia et al., 2021). Yet, it is unclear whether DNAm sex differences are due to different biological pathways, socialisation, or both (Gutman & Codiroli McMaster, 2020).

### Strengths and limitations

This investigation of DNAm and internalising problems in very young children complements existing evidence, which mainly comprises adult populations (Barker et al., 2018). The inclusion of two different DNAm assessment time points (cord- and peripheral-blood), enabled capturing prenatal and postnatal risk factors. The investigation of internalising problems at age 3 and age 6 allowed evaluating the persistence of these effects across development. Another strength is in the application of an epigenome-wide probe-level and regional analysis within a multi-cohort framework, in a hypothesis-free way, which may reveal novel biological pathways to anxiety and depression. Last, the relatively large sample size and prospective design helped indicate the temporal sequence of events and reduced recall bias.

The results of this study should be interpreted considering the following limitations. Statistical power was compromised by the high zero inflation of children’s internalising problems at both time-points. The demographic profile suggests children were likely at low risk for adverse experiences and thus it is unclear whether results generalize to less advantaged populations. Selection bias is common in longitudinal cohort studies (Boyd et al., 2013; Taylor et al., 2018) and families with a less advantaged background and more mental health problems are more likely to drop-out (Magnus et al., 2016; Wolke et al., 2009). This bias may be stronger for the DNAm subsamples, which are commonly selected based on completeness of other cohort variables. The database search may be influenced by the research focus on certain phenotypes, such as schizophrenia, which has been comprehensively studied in psychiatric genetics (Zakaria et al., 2023). Last, this study relied on DNAm assessed in blood, yet the tissue of most interest for mental health phenotypes is brain tissue. There is some evidence that part of blood DNAm can function as a proxy for brain DNAm (Davies et al., 2012; Kaminsky et al., 2009; Walton et al., 2016). Regardless of tissue concordance, these results may contribute to developing DNAm biomarkers for early detection of internalising problems (Walton et al., 2019).

### Future research

Future EWAS with larger sample sizes are needed to clarify the relationship between DNAm and internalising problems. The higher-powered regional analysis found more evidence for associations between DNAm and internalising problems, warranting replication by independent studies. Future research using methylation profile scores could complement this study’s approach by leveraging subtle changes in DNAm across the genome (Thompson et al., 2022). Studies should invest in more detailed phenotyping of anxiety and depression in children (Dunn et al., 2019), include multiple assessment time-points, and ideally draw on reports from multiple informants (Sallis et al., 2019). Also neuroimaging phenotypes may be included, which may serve as intermediate phenotypes for mental health problems (Walton et al., 2023). Methods like longitudinal mixed models or Structured Life Course Modeling Approach could help to capture developmental trajectories. Replication in more diverse populations with higher risks for adverse early life experiences is necessary and more extensive research is needed to understand the observed sex differences and their clinical significance. Investigation of gender factors, such as caregiver expectations and gendered stress, could help understand how sociocultural factors shape DNA methylation and mental health (Cattaneo et al., 2024). Future research might explore genotype-DNAm interactions, as certain genotypes may be more susceptible to environmental effects (Brummelte et al., 2017; Oberlander et al., 2010; Olsson et al., 2010; Teh et al., 2014). Additionally, future studies should shift focus from early adverse experiences to exploring resilience factors in children, examining how positive early-life experiences affect DNAm and mental health (Kentner et al., 2019).

## CONCLUSION

The findings of this study provide some evidence for associations between DNAm and childhood internalising problems, with distinct signals observed at different assessment time points for both DNAm (birth vs. childhood) and internalising problems (at ages 3 and 6). Despite the high zero inflation of internalizing problems, the DMR analyses suggested differences in DNAm at regions annotated to genes, which have previously been associated with mental health phenotypes related to internalising problems. It is likely that these effects would be further exacerbated in populations with more occurrences of internalising problems. As anxiety and depression are highly polygenic disorders, which are likely influenced by the interplay of small effects of many different genes, more EWAS are needed to replicate or invalidate findings of this study, as well as to detect novel differentially methylated regions, which might have been missed.

## ETHICAL INFORMATION

Ethical approval was obtained by local ethics committees and informed consent for the use of data was obtained from all participants (Supporting Information File 1).

## Data Availability

All data produced in the present study are available upon reasonable request to the authors

https://ewascatalog.org/

https://www.ebi.ac.uk/gwas/

https://www.genecards.org/

---

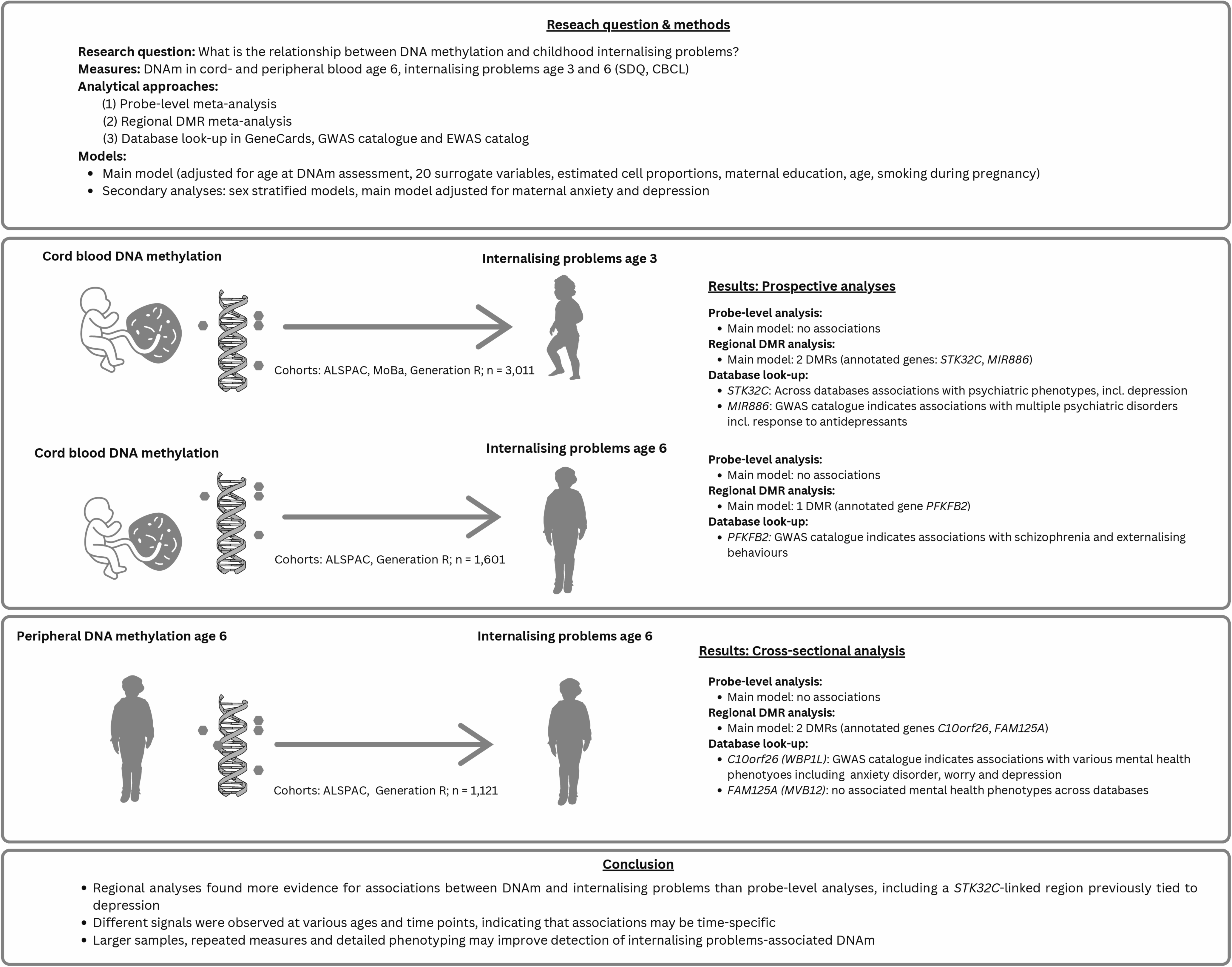

## Notes

Funding statement: This work was supported by multiple funders. Detailed funding information for each participating cohort is provided in the Supporting Information File S1.

### Competing Interest Statement

The authors have declared no competing interest.

### Funding Statement

This study was funded by multiple funders. Please refer to Supporting Information File S1 for an overview of all funders.

### Author Declarations

Ethical approval was obtained for each cohort by local ethics committees and informed consent for the use of data was obtained from all participants. For further information see Supporting Information File S1

