## Supporting Information File S1 for "The development of childhood internalising problems: A meta-analysis of epigenome-wide-association-studies"

### Avon Longitudinal Study of Parents and Children (ALSPAC)

#### Description of the cohort

Ethics approval for the study was obtained from the ALSPAC Ethics and Law Committee and the Local Research Ethics Committees. Informed consent for the use of data collected via questionnaires and clinics was obtained from participants following the recommendations of the ALSPAC Ethics and Law Committee at the time.

Pregnant women resident in Avon, UK with expected dates of delivery between 1st April 1991 and 31st December 1992 were invited to take part in the study. 20,248 pregnancies have been identified as being eligible and the initial number of pregnancies enrolled was 14,541. Of the initial pregnancies, there was a total of 14,676 foetuses, resulting in 14,062 live births and 13,988 children who were alive at 1 year of age. When the oldest children were approximately 7 years of age, an attempt was made to bolster the initial sample with eligible cases who had failed to join the study originally. As a result, when considering variables collected from the age of seven onwards (and potentially abstracted from obstetric notes) there are data available for more than the 14,541 pregnancies mentioned above: The number of new pregnancies not in the initial sample (known as Phase I enrolment) that are currently represented in the released data and reflecting enrolment status at the age of 24 is 906, resulting in an additional 913 children being enrolled (456, 262 and 195 recruited during Phases II, III and IV respectively). The phases of enrolment are described in more detail in the cohort profile paper and its update (see footnote 5 below). The total sample size for analyses using any data collected after the age of seven is therefore 15,447 pregnancies, resulting in 15,658 foetuses. Of these 14,901 children were alive at 1 year of age. Of the original 14,541 initial pregnancies, 338 were from a woman who had already enrolled with a previous pregnancy, meaning 14,203 unique mothers were initially enrolled in the study. As a result of the additional phases of recruitment, a further 630 women who did not enrol originally have provided data since their child was 7 years of age. This provides a total of 14,833 unique women (G0 mothers) enrolled in ALSPAC as of September 2021. G0 partners were invited to complete questionnaires by the mothers at the start of the study and they were not formally enrolled at that time. 12,113 G0 partners have been in contact with the study by providing data and/or formally enrolling when this started in 2010. 3,807 G0 partners are currently enrolled.

Please note that the study website contains details of all the data that is available through a fully searchable data dictionary and variable search tool:

<http://www.bristol.ac.uk/alspac/researchers/our-data/>

#### DNA methylation data

As part of the Accessible Resources for Integrated Epigenomic Studies (ARIES, http://www.ariesepigenomics.org.uk/) project, DNA methylation was generated for 1018 mother-offspring pairs from the ALSPAC cohort, using the Infinium HumanMethylation450 BeadChip array (Illumina Inc., San Diego, United States). ARIES participants were selected based on availability of DNA samples at two time points for the mother (antenatal and at follow-up when the offspring were adolescents) and at three time points for the offspring (neonatal, childhood (age 7), and adolescence (age 17)). The current study used child cord blood at birth and whole blood at age 7. Consent for biological samples has been collected in accordance with the Human Tissue Act (2004).

Methods for methylation measurements in ALSPAC have been described previously (Relton et al., 2015). Briefly, cord blood was collected according to standard procedures. DNA methylation assays and data pre-processing was performed at the University of Bristol as part of the ARIES project. DNA was extracted using standard protocol and was bisulfite-converted using the Zymo EZ DNA MethylationTM kit (Zymo, Irvine, CA). DNA methylation was then measured using the Infinium HM450 BeadChip assay (Illumina Inc, San Diego, CA), according to the standard protocol. Arrays were scanned using an Illumina iScan. An initial review of data quality was assessed using GenomeStudio (version 2011.1). A semi-random approach (sampling criteria were in place to ensure that all time points were represented on each array) was used to distribute ARIES samples across slides to minimize the possibility of potential confounding by batch. Data were normalised using the meffil R package (Min et al, 2018) using the functional normalisation approach.

#### Definition of variables and covariates

Offspring internalising problems: In ALSPAC, internalising problems at the age 3 and 7 were assessed through questionnaires at 47 months and 81 months, respectively. The total emotional symptoms score was calculated by summing the score of each item for children that had complete data on all 5 items. To obtain the item mean, the total score was divided by 5 (the number of items of the emotional symptoms score)

Offspring age: Offspring’s age at completion of the questionnaires was first generated based on offspring’s date of birth and the date of completion of the questionnaires. Offspring’s age was transformed from days to weeks by dividing age in days by 7.

Maternal education (as proxy for maternal socioeconomic position): Maternal education was assessed in week 32 of gestation and coded as an ordinal variable: "Vocational/CSE" = 1, “O level” (at 16, equivalent to lower grades of ordinary-level) = 2, "A level" (ordinary-level school-leaving certificate (at 18) = 3, and “Degree” (advanced-level school-leaving certificate (post-18)/degree) = 4.

Maternal smoking during pregnancy was assessed as an ordinary variable representing 0 = no or early smoking during pregnancy, 1 = Stopped before the second trimester of pregnancy and 2 = Smoking in the third trimester or throughout pregnancy.

Maternal age continuous numeric variable in years assessed at birth of study child.

Parity has been assessed at 18 weeks gestation as number of previous pregnancies resulting in either a livebirth or a stillbirth.

Gestational age was calculated (in days) based on the date of the mother’s last menstrual period (LMP) when the mother was certain of this, but for uncertain LMPs and conflicts with clinical assessment the ultrasound assessment was used. Where maternal report and ultrasound assessment conflicted, an experienced obstetrician reviewed clinical records and made a best estimate.

Maternal anxiety and depression during pregnancy: Maternal anxiety and depressive symptoms during pregnancy were assessed with the Edinburgh Postnatal Depression scale in a questionnaire at 18-weeks gestation. The questionnaire consists of 10 items rated on 4-point scale ranging from 0 = “Not at all” to 3 = “yes, most of the time”) (Cox et al., 1987). A total score was generated if mothers had data on each of the ten items by summing up the individual item scores.

Offspring sex was taken from obstetric records.

### MoBa

#### Description of the cohort

The Norwegian Mother, Father and Child Cohort Study (MoBa) is a population-based pregnancy cohort study conducted by the Norwegian Institute of Public Health. Participants were recruited from all over Norway from 1999-2008 (Magnus et al. 2016; <https://www.fhi.no/en/studies/moba/>). The women consented to participation in 41% of the pregnancies. The cohort includes approximately 114.500 children, 95.200 mothers and 75.200 fathers.

The establishment of MoBa and initial data collection was based on a license from the Norwegian Data Protection Agency and approval from The Regional Committees for Medical and Health Research Ethics. The MoBa cohort is currently regulated by the Norwegian Health Registry Act. The current study was approved by The Regional Committees for Medical and Health Research Ethics

MoBa1 and MoBa2 are subsets of a larger study within MoBa that included a cohort random sample and cases of asthma at age three years (Haberg et al., 2011). These data set are well described in earlier papers by the PACE consortia (Joubert et al., 2012; Küpers et al., 2019). Years of birth were 2002-2004 for children in MoBa1 and 2000-2005 for MoBa2.

The establishment of MoBa and initial data collection was based on a license from the Norwegian Data Protection Agency and approval from The Regional Committees for Medical and Health Research Ethics. MoBa is currently regulated by the Norwegian Health Registry Act. The study was approved by the Regional Committee for Ethics in Medical Research, Norway (#2017/1362). In addition, MoBa1 was approved by the Institutional Review Board of the National Institute of Environmental Health Sciences, USA. The consent given by the participants does not allow for storage of data on an individual level in repositories or journals. Researchers who want access to data sets for replication should submit an application to <http://www.helsedata.no/>. Access to data sets requires approval from The Regional Committee for Medical Research Ethics in Norway and an agreement with MoBa.

#### DNA methylation data

Details of the DNA methylation measurements and quality control for the MoBa1 participants were previously described (Joubert et al., 2012) and the same protocol was implemented for the MoBa2 participants. Briefly, umbilical cord blood samples were collected and frozen at birth at -80°C. All biological material was obtained from the Biobank of the MoBa study (Paltiel et al., 2014). Bisulfite conversion was performed using the EZ-96 DNA Methylation kit (Zymo Research Corporation, Irvine, CA) and DNA methylation was measured at 485,577 CpGs in cord blood using Illumina’s Infinium HumanMethylation450 BeadChip. Raw intensity (.idat) files were handled in R using the minfi package to calculate the methylation level at each CpG as the beta-value (β=intensity of the methylated allele (M)/(intensity of the unmethylated allele (U) + intensity of the methylated allele (M) + 100)) and the data was exported for quality control and processing. Control probes (N=65) and probes on X (N=11 230) and Y (N=416) chromosomes were excluded in both datasets. Remaining CpGs missing > 10% of methylation data were also removed (N=20). Samples indicated by Illumina to have failed or have an average detection p value across all probes < 0.05 (N=49) and samples with gender mismatch (N=13 MoBa1) were also removed. We accounted for the two different probe designs by applying the intra-array normalization strategy Beta Mixture Quantile dilation (BMIQ). After quality control exclusions, the sample size for MoBa1 was 1,068 and 685 for MoBa2.

#### Definition of variables and covariates

Offspring internalising problems: Internalising problems at the age 3 were assessed through a questionnaire at 36 months. Nine selected items of the original CBCL internalising subscale (including somatic complaints) were used to calculate a total internalising problems score by summing the score of each item for children that had complete data on all 9 items.

Offspring age: Offspring’s age at completion of the questionnaires was generated based on offspring’s date of birth and the date of completion of the questionnaires.

Maternal education (as proxy for maternal socioeconomic position): Maternal education was assessed at 15 weeks gestation and coded as an ordinal variable: "<High Sch " = 0, “High School degree” = 1, " Some college " = 2, and “+4yr College” = 3.

Maternal smoking during pregnancy was assessed as an ordinary variable representing 0 = no or early smoking during pregnancy, 1 = Stopped before the second trimester of pregnancy and 2 = Smoking in the third trimester or throughout pregnancy.

Maternal age continuous numeric variable in years collected from Medical Birth Registry of Norway (MBRN), a national health registry containing information about all births in Norway.

Parity was recorded in MBRN.

Gestational age was recorded in MBRN, using ultrasound measurement when available, and LMP otherwise.

Maternal anxiety and depression during pregnancy: Maternal anxiety and depressive symptoms during pregnancy were assessed with five items from the (Hopkins) Symptoms Checklist-25 (SCL-25) (Tambs & Moum, 1993): the SCL-5 (Tambs & Røysamb, 2014). Items from the SCL-25 such as feeling constantly frightened or anxious, worrying too much, and feeling blue are scored on a scale ranging from 1 (“not bothered”) to 4 (“very bothered”). The SCL-5 has been validated with a correlation of 0.92 with the SCL-25 (Tambs & Røysamb, 2014). Maternal anxiety and depression were assessed at 15 weeks gestation using the SCL-5.

Mothers reported on their anxiety and depressive symptoms in a questionnaire at gestational week 15.

Offspring’s sex was recorded in the MBRN.

**Acknowledgement**:

The Norwegian Mother, Father and Child Cohort Study is supported by the Norwegian Ministry of Health and Care Services and the Ministry of Education and Research. We are grateful to all the participating families in Norway who take part in this on-going cohort study.

### Generation R

#### Description of the cohort

The Generation R Study (<https://generationr.nl/researchers/>) is a population-based prospective cohort study from fetal life onwards in Rotterdam, the Netherlands. A detailed description can be found elsewhere [Kooijman et al, Eur J Epidemiol 2016; Kruithof et al, Eur J Epidemiol 2014]. Briefly, 9,778 pregnant women with a delivery date between April 2002 and January 2006 were included (response rate at baseline 61%). There is ongoing follow-up. The study was approved by Medical Ethical Committee of Erasmus MC, University Medical Center Rotterdam and written consent was obtained for all participants. For this analysis, we used singleton live births with epigenome-wide association arrays and information on paternal BMI and complete covariates, giving total sample sizes of 947 at birth and 335 in childhood.

#### DNA methylation data

DNA, 500 ng per sample, extracted (using the salting-out method) from blood samples taken at birth (cord blood) or at the 5-year follow-up underwent bisulfite conversion using the EZ-96 DNA Methylation kit (Shallow) (Zymo Research Corporation, Irvine, USA). Samples were plated onto 96-well plates in no specific order. Samples were processed with the Illumina Infinium HumanMethylation450 BeadChip (Illumina Inc., San Diego, USA), which analyses methylation at 485,577 CpG sites. Preparation and normalization of the HumanMethylation450 BeadChip array data was performed according to the CPACOR workflow [Lehne et al. 2015] using the package minfi in R [R Core Team, 2013]. Probes that had a detection p-value above background (based on sum of methylated and unmethylated intensity values) ≥ 1E-16 were set to missing per array. Next, the intensity values were stratified by autosomal and non-autosomal probes and quantile normalized for each of the six probe type categories separately: type II red/green, type I methylated red/green and type I unmethylated red/green. Beta values were calculated as proportion of methylated intensity value on the sum of methylated+unmethylated+100 intensities. Arrays with observed technical problems such as failed bisulfite conversion, hybridization or extension, as well as arrays with a mismatch between sex of the proband and sex determined by the chr X and Y probe intensities were removed from subsequent analyses. Additionally, only arrays with a call rate > 95% per sample were processed further.

#### Definitions of variables and covariates

Offspring internalising problems: Internalising problems at the age 3 and 7 were assessed through questionnaires at 36 months and 72 months, respectively. The CBCL internalising scale score was generated according to the CBCL manual.

Offspring age at internalising problems assessment was reported by mothers in the corresponding questionnaire.

Maternal education (as proxy for maternal socioeconomic position): Maternal education was assessed in week 12-20 of gestation and coded as an ordinal variable: " “no education finished " = 1, “primary” (at 12, equivalent to lower grades of ordinary-level) = 2, "secondary, phase 1" (ordinary-level school-leaving certificate (at 16) = 3, and “3-secondary, phase 2” (advanced-level school-leaving certificate (post-16)) = 4, “higher, phase 2” (education after High School) = 5.

Maternal smoking during pregnancy was assessed as an ordinary variable representing 0 = no or early smoking during pregnancy, 1 = Stopped before the second trimester of pregnancy and 2 = Smoking in the third trimester or throughout pregnancy.

Maternal age continuous numeric variable in years assessed at study intake.

Gestational age was assessed at birth of study child.

Maternal anxiety and depression during pregnancy was assessed through questionnaires at weeks 20-25 of gestation using the Brief Symptom Inventory (12 items, rated on a 5-point scale ranging from 0 = “Not at all” to 4 = “extremely”) (Derogatis & Melisaratos, 1983).

Offspring sex was derived from birth records.

### Methods

Mothers reported on their children’s internalising problems over the last two months, on a 3-point scale, ranging from 0 = “Not True” to 2 = “Very True or Often True”, with higher scores reflecting more internalising problems.

MoBa1 and MoBa2 used a short scale, which contained a subset of 9-items of the full CBCL internalising scale. Information on the selection of items can be found here <https://www.fhi.no/globalassets/dokumenterfiler/studier/den-norske-mor-far-og-barn--undersokelsenmoba/instrumentdokumentasjon/instrument-documentation-q6.pdf>).

The emotional symptoms scale of the SDQ incorporates 5 items that are rated by mothers on a 3-point scale (0 = Not true, 1 = Somewhat true, 2 = Certainly true), with higher scores reflecting more internalising problems. Mothers were asked to report on their child’s behaviours over the past six months.

Comparability of the questionnaires. Previous research from the UK (Goodman & Scott, 1999), Germany, and Austria (Klasen, 2000) has shown that the internalising problems subscale of the CBCL and emotional symptoms subscale of the SDQ correlate moderately to highly (r = 0.69 - 0.74) in 4 to 10-year-old children and can equally well distinguish between clinical and control populations. The comparability of the scales is further supported by studies from the Netherlands (van Widenfelt et al., 2003) and Finland (Koskelainen & Kaljonen, 2000), which found the CBCL and SDQ to be comparable in samples of older children aged 11-16 years.

Offspring age. Due to established associations between age and variation in DNA methylation (Teschendorff, West, et al., 2013), the cord blood analyses were adjusted for gestational age in days and the peripheral blood analyses for offspring’s age at DNA methylation assessment.

Maternal education. An ordinal variable of maternal education, with higher scores representing a higher level of education, was used as a proxy for family socioeconomic position (SEP). Family SEP has been found to be associated with higher risk for offspring mental health problems (Barker et al., 2012; Goodman et al., 2011) and is commonly adjusted for in EWAS (Joubert et al., 2016; Sharp et al., 2021).

Maternal age (years). Maternal age was found to be associated with both offspring mental health problems (Carslake et al., 2017), and DNA methylation (Markunas et al., 2016).

Maternal smoking during pregnancy. Maternal smoking during pregnancy was included as an ordinal variable (0 = no smoking, 1 = giving up smoking early in pregnancy, 2 = smoking throughout pregnancy) and included because of the strong influence on DNA methylation (Joubert et al., 2016) and indication for association with offspring internalising problems (Ashford et al., 2008b; Moylan et al., 2015).

### Results

#### Quality control checks

##### Cohort results

In each cohort, the effect estimates of each of the models correlated moderately to highly and the pattern of correlations was consistent across cohorts, assessment time-points, and tissue of DNA methylation (cord vs. peripheral blood), indicating that each model was assessing the same effect (Figures S1 to S8). Lowest correlations were observed between the effect estimates of the sex-stratified models, which may be explained by the small sample size in the stratified analyses, residual confounding (Yousefi et al., 2015), or sex-differences between the associations of DNA methylation and internalising problems. As most of the *p*-values in the QQ-plots follow the null line closely and stay within the 95% Confidence Intervals, there is little evidence for systematic confounding of the association between DNA methylation at individual CpG sites and offspring internalising problems (Figure S9 to S16).

Figure S1
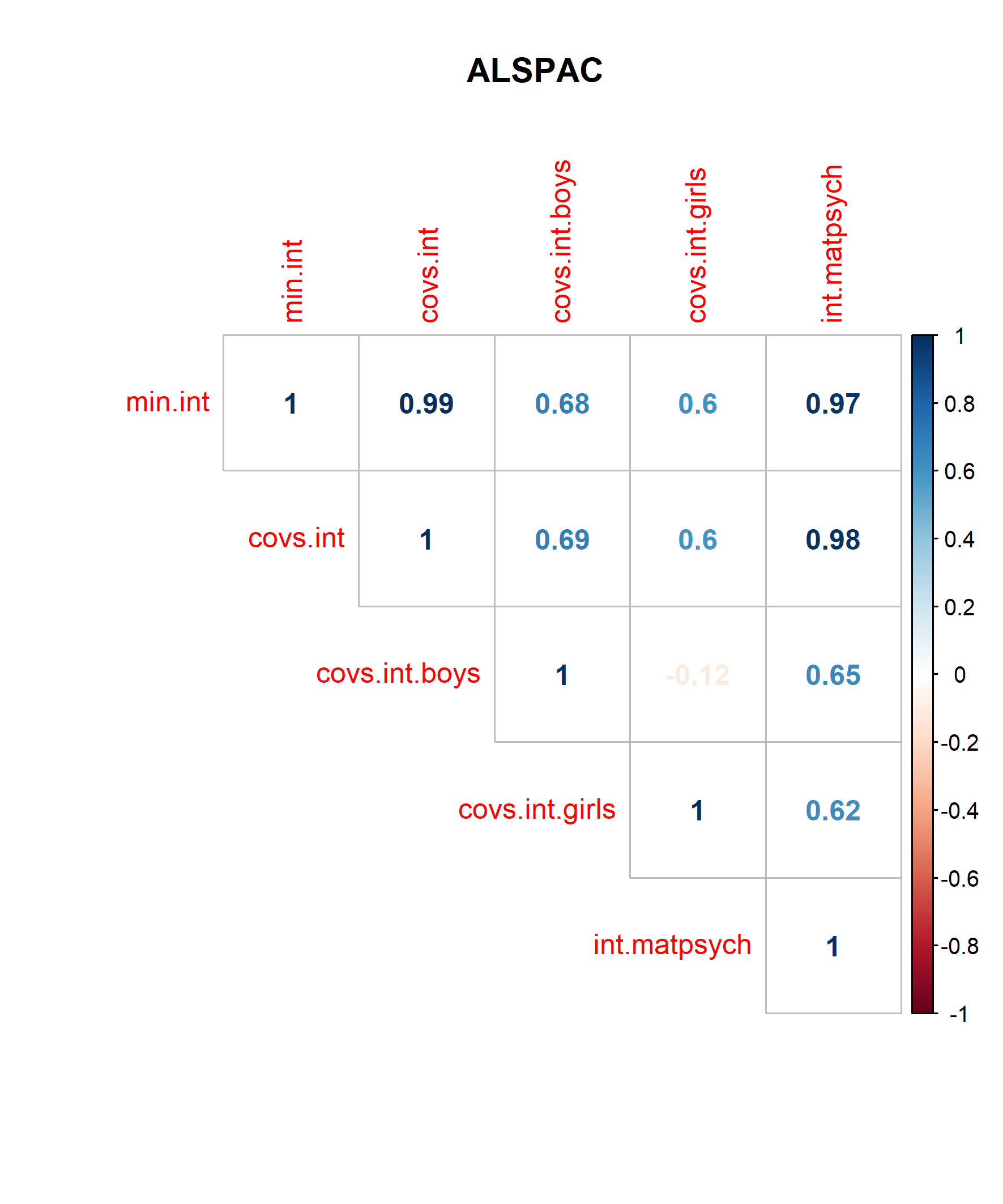
Correlation plot cord blood analysis age 3 – ALSPAC
Figure S2 Correlation plot cord blood analysis age 3 – Generation
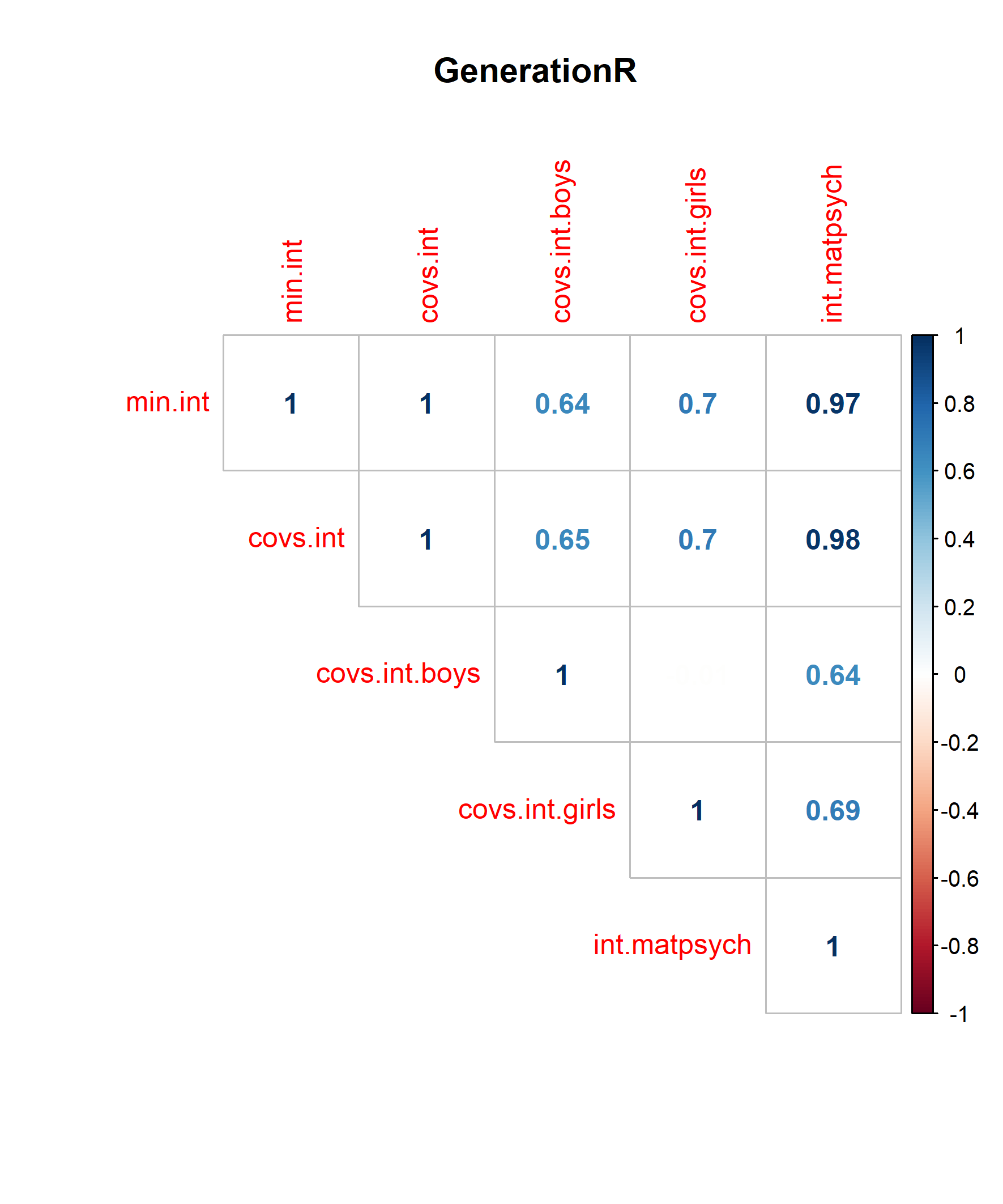

Figure S3
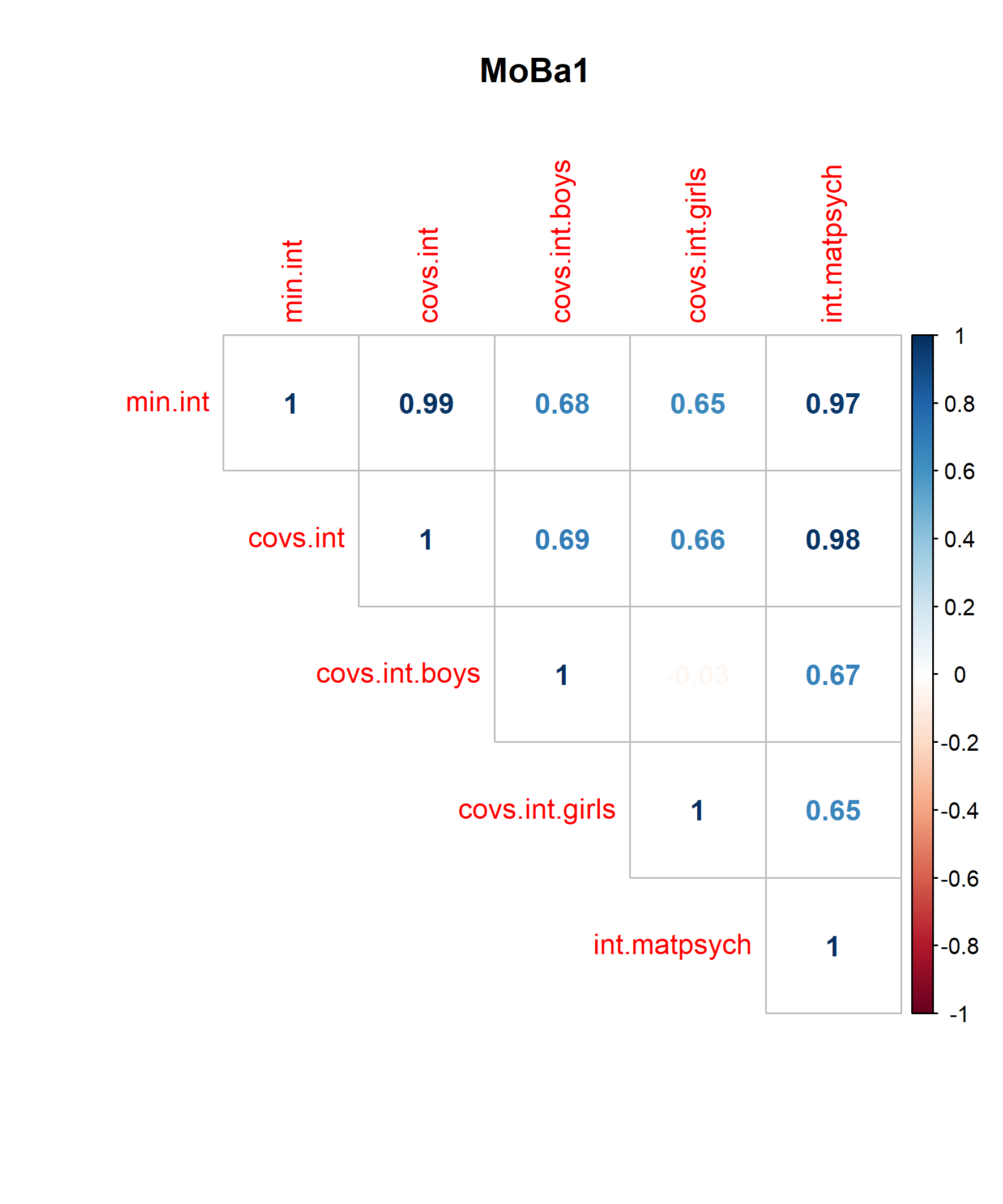
Correlation plot cord blood analysis age 3 – MoBa1

Figure S4 Correlation plot cord blood analysis age 3 – MoBa2

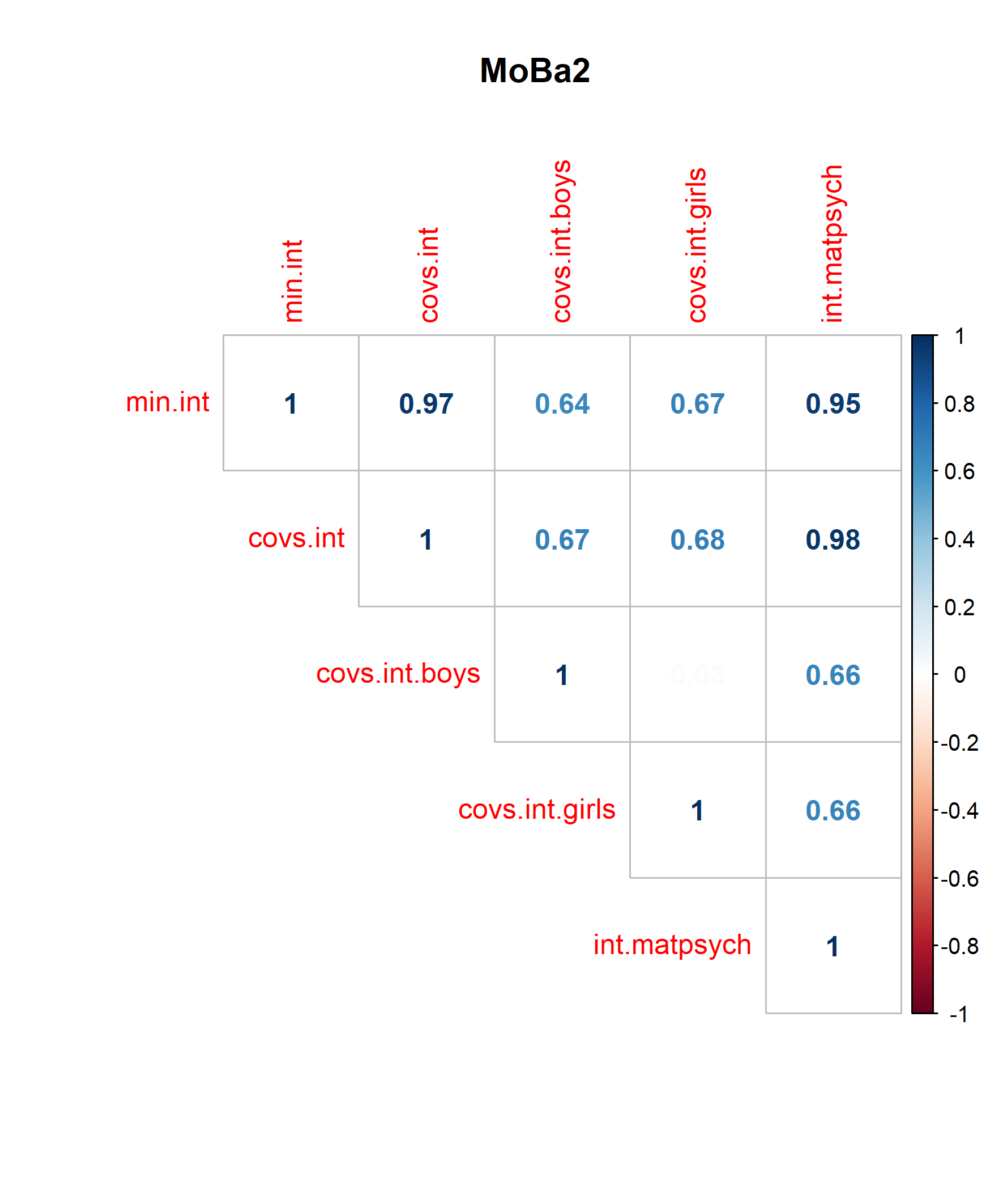

Figure S5 Correlation plot cord blood analysis age 7 – ALSPAC
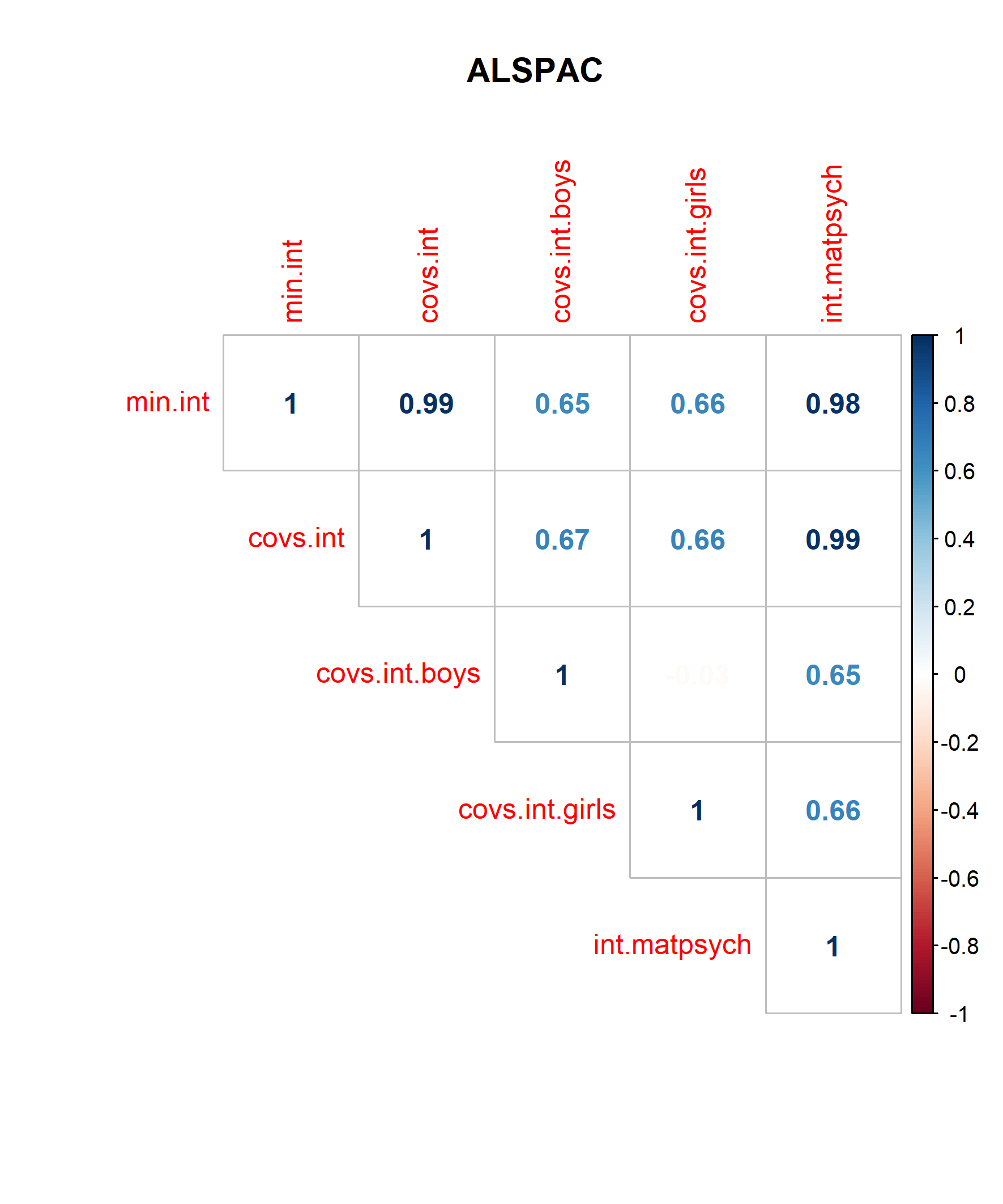

Figure S6 Correlation plot cord blood analysis age 7 – Generation R
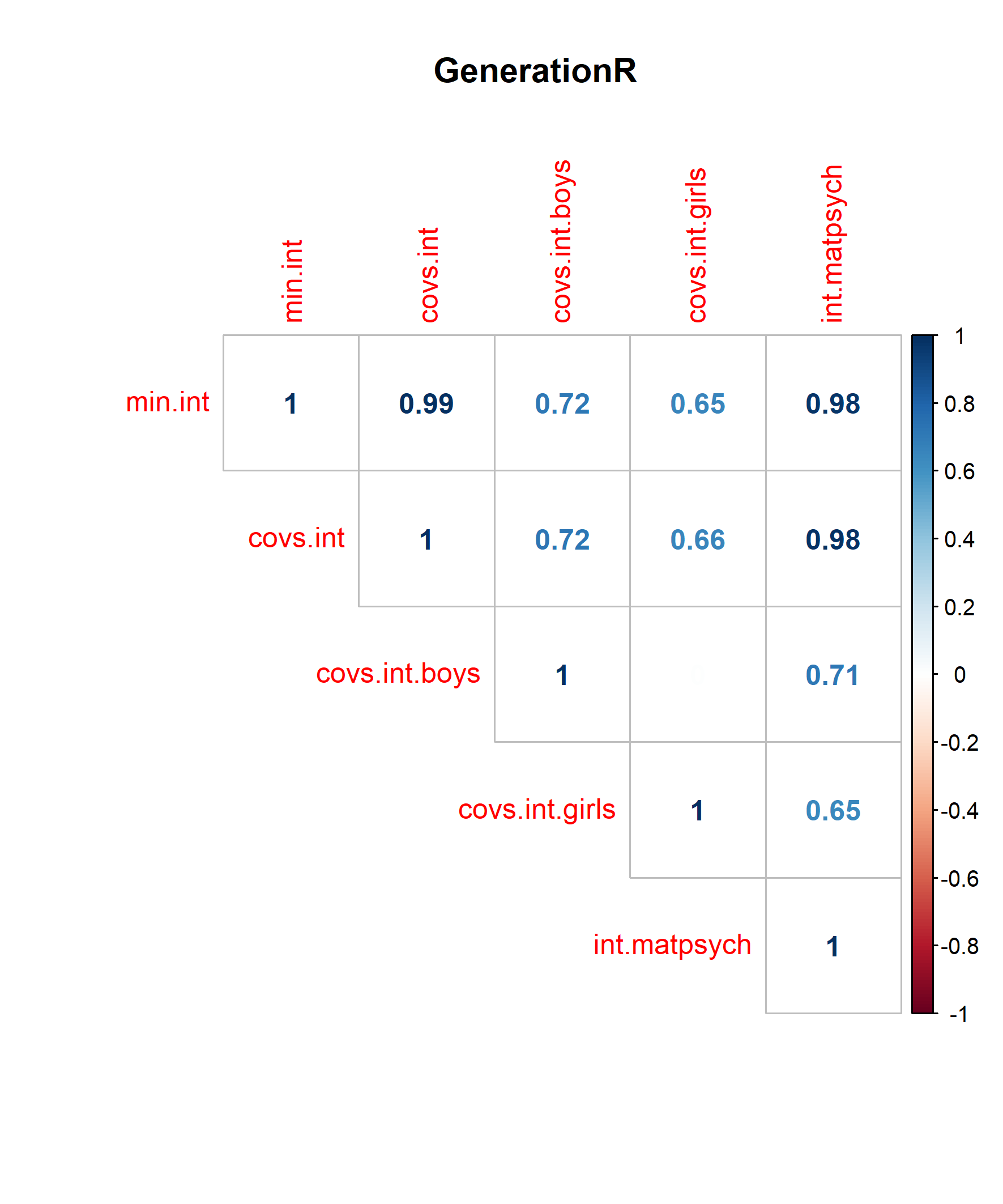

Figure S7 Correlation plot cross-sectional -analysis age 7 – ALSPAC

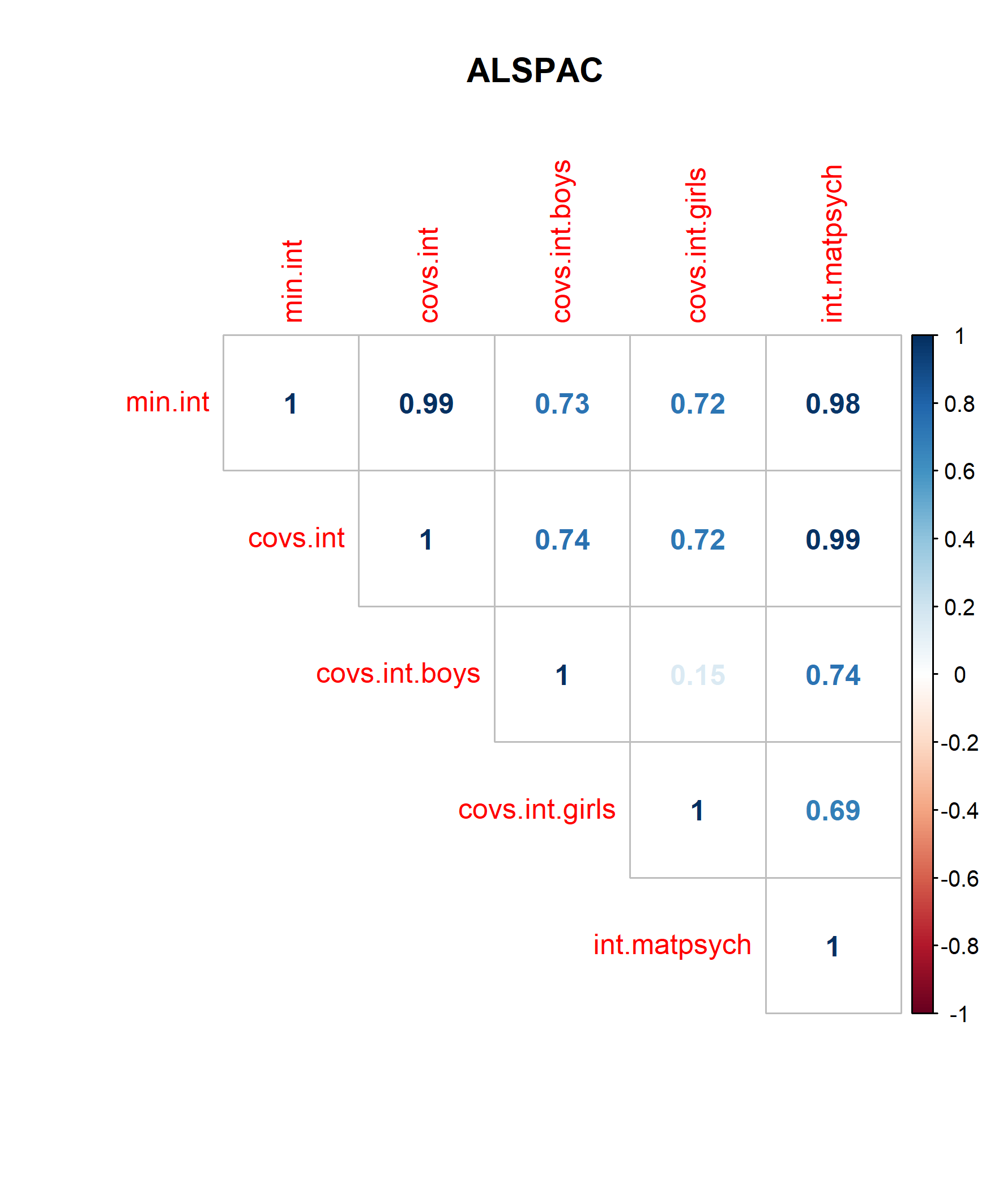

Figure S8 Correlation plot cross-sectional analysis age 7 – Generation R

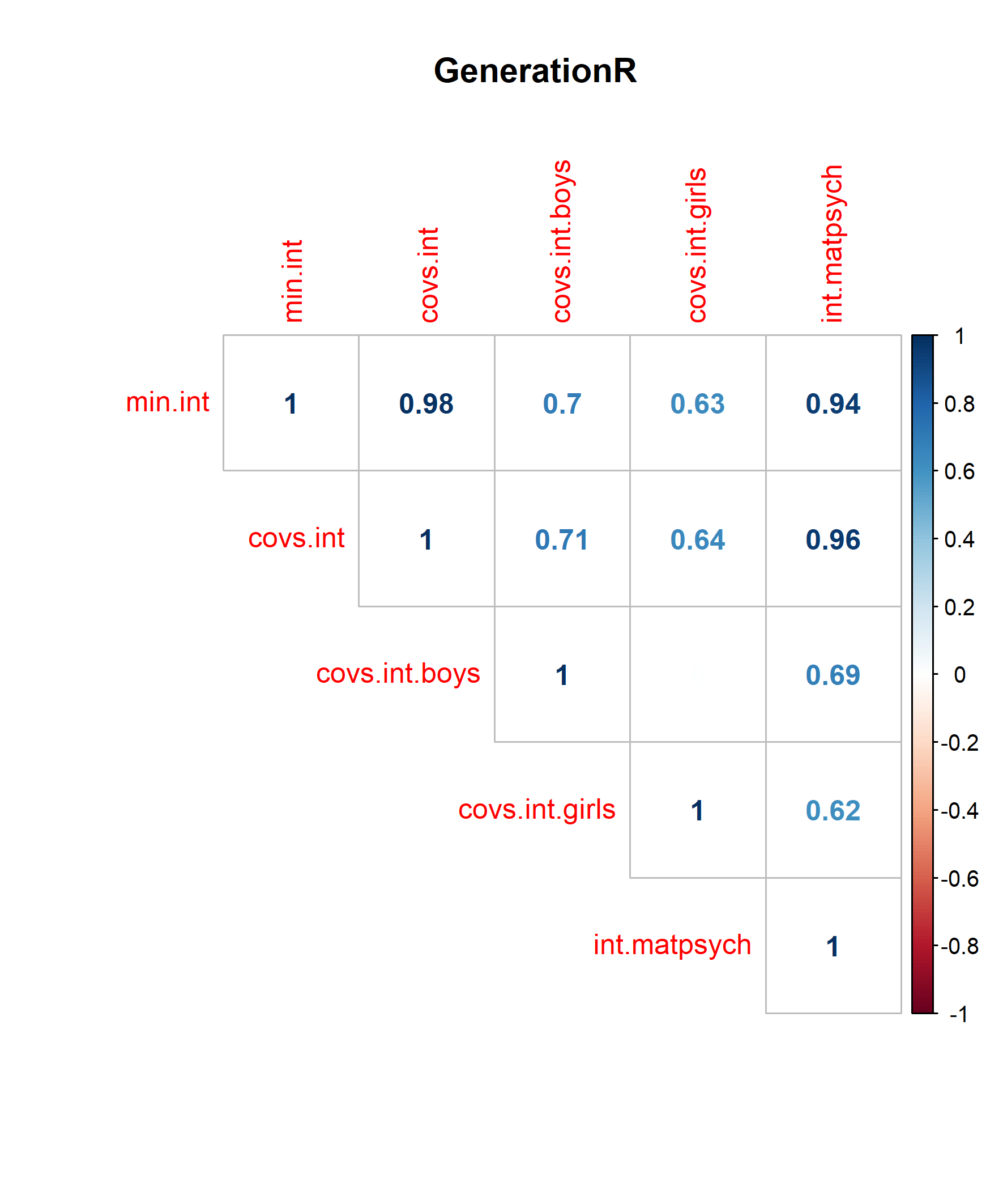

Figure S9 QQ-plots cord blood analysis age 3 – ALSPAC
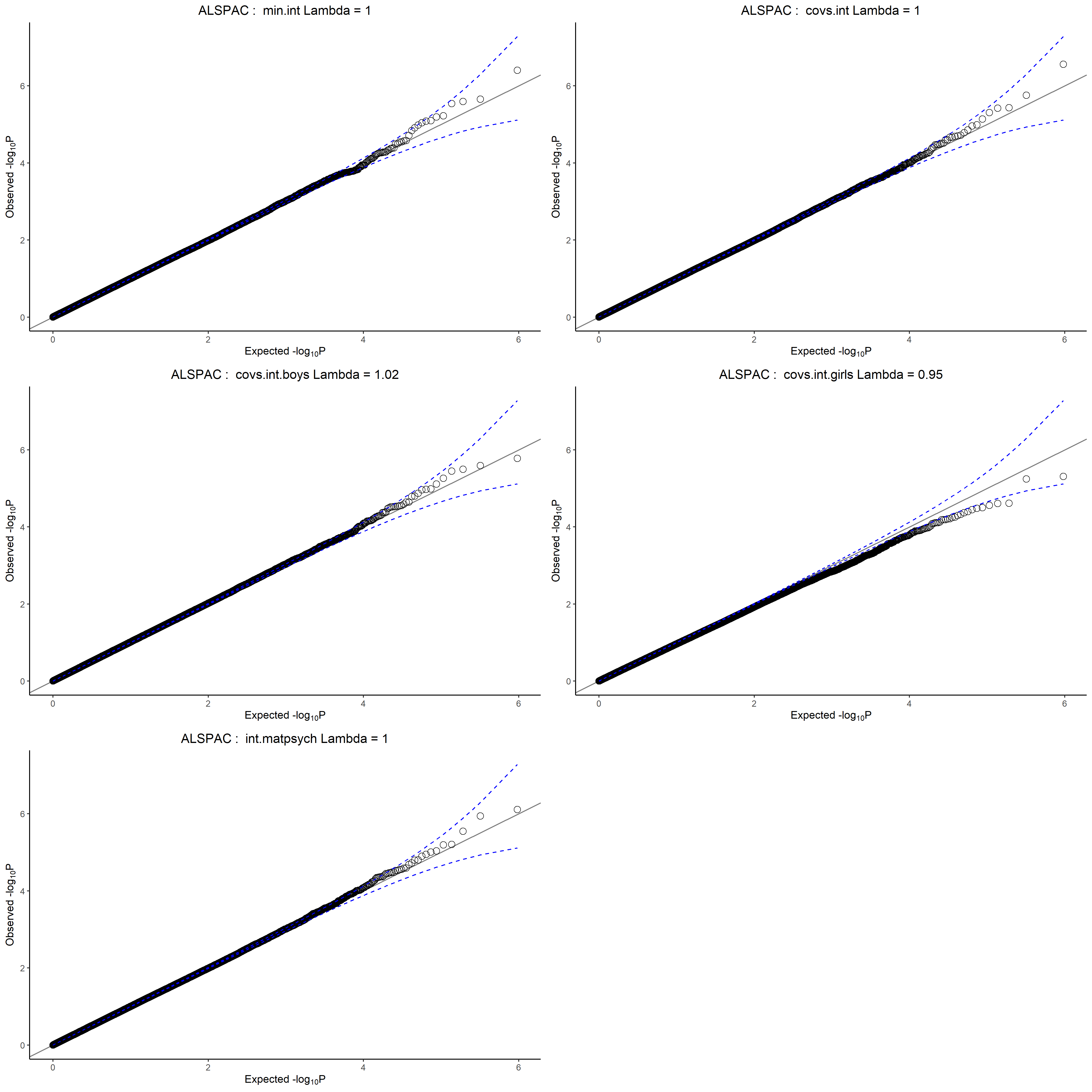

Note. Quantile-Quantile (Q-Q) plot of the DNAm signals across the different models. min.int = minimally adjusted model, covs.int = covariate adjusted model, covs.int.boys = covariate, male-sex stratified model, covs.int.girls = covariate, female-sex stratified model, int.matpsych = maternal anxiety/depression adjusted model.

Figure S10 QQ-plots cord blood analysis age 3 – Generation R
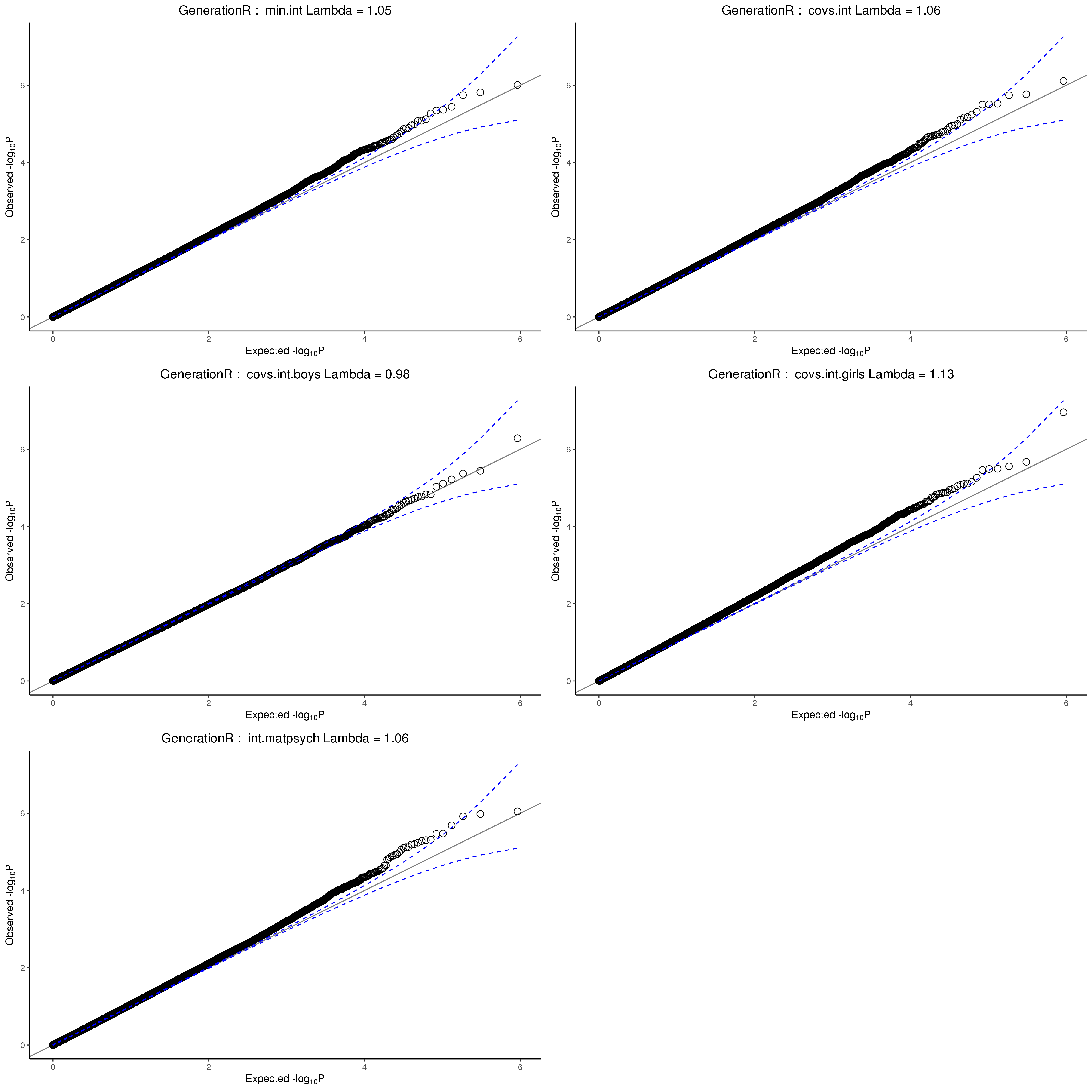

Note. Quantile-Quantile (Q-Q) plot of the DNAm signals across the different models. min.int = minimally adjusted model, covs.int = covariate adjusted model, covs.int.boys = covariate, male-sex stratified model, covs.int.girls = covariate, female-sex stratified model, int.matpsych = maternal anxiety/depression adjusted model.

Figure S11 QQ-plots cord blood analysis age 3 – MoBa1

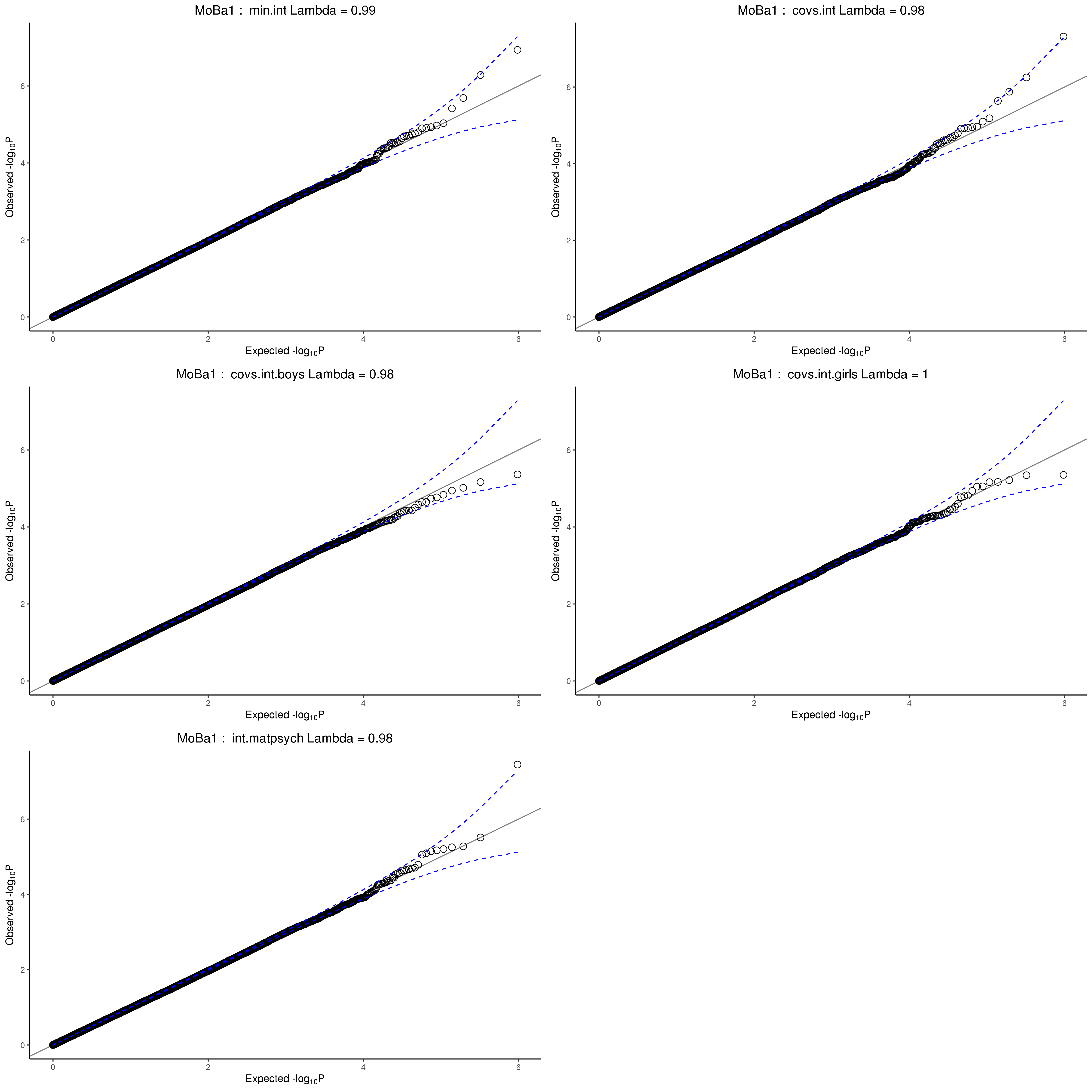

Note. Quantile-Quantile (Q-Q) plot of the DNAm signals across the different models. min.int = minimally adjusted model, covs.int = covariate adjusted model, covs.int.boys = covariate, male-sex stratified model, covs.int.girls = covariate, female-sex stratified model, int.matpsych = maternal anxiety/depression adjusted model.

Figure S12 QQ-plots cord blood analysis age 3 - MoBa2

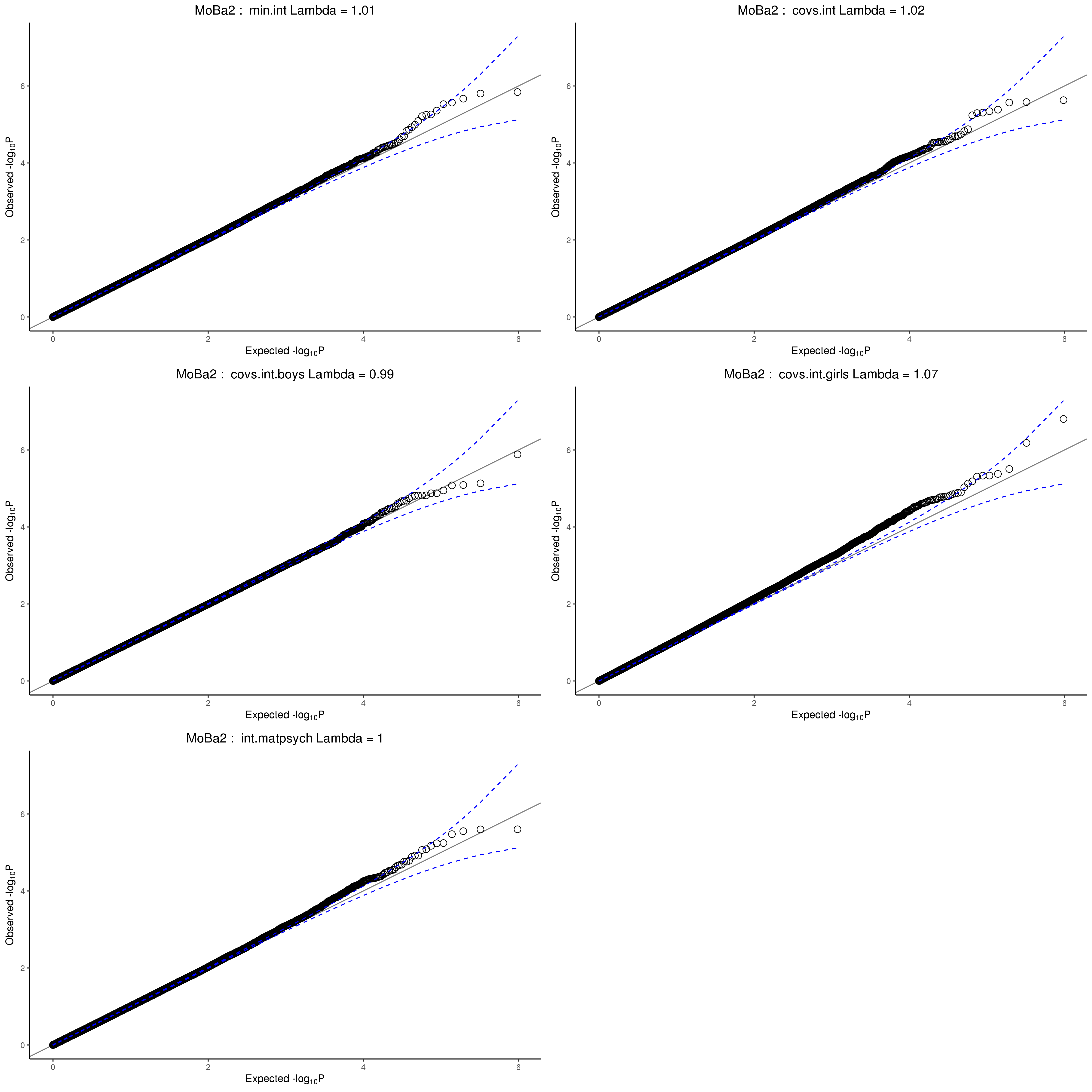

Note. Quantile-Quantile (Q-Q) plot of the DNAm signals across the different models. min.int = minimally adjusted model, covs.int = covariate adjusted model, covs.int.boys = covariate, male-sex stratified model, covs.int.girls = covariate, female-sex stratified model, int.matpsych = maternal anxiety/depression adjusted model.

Figure S13
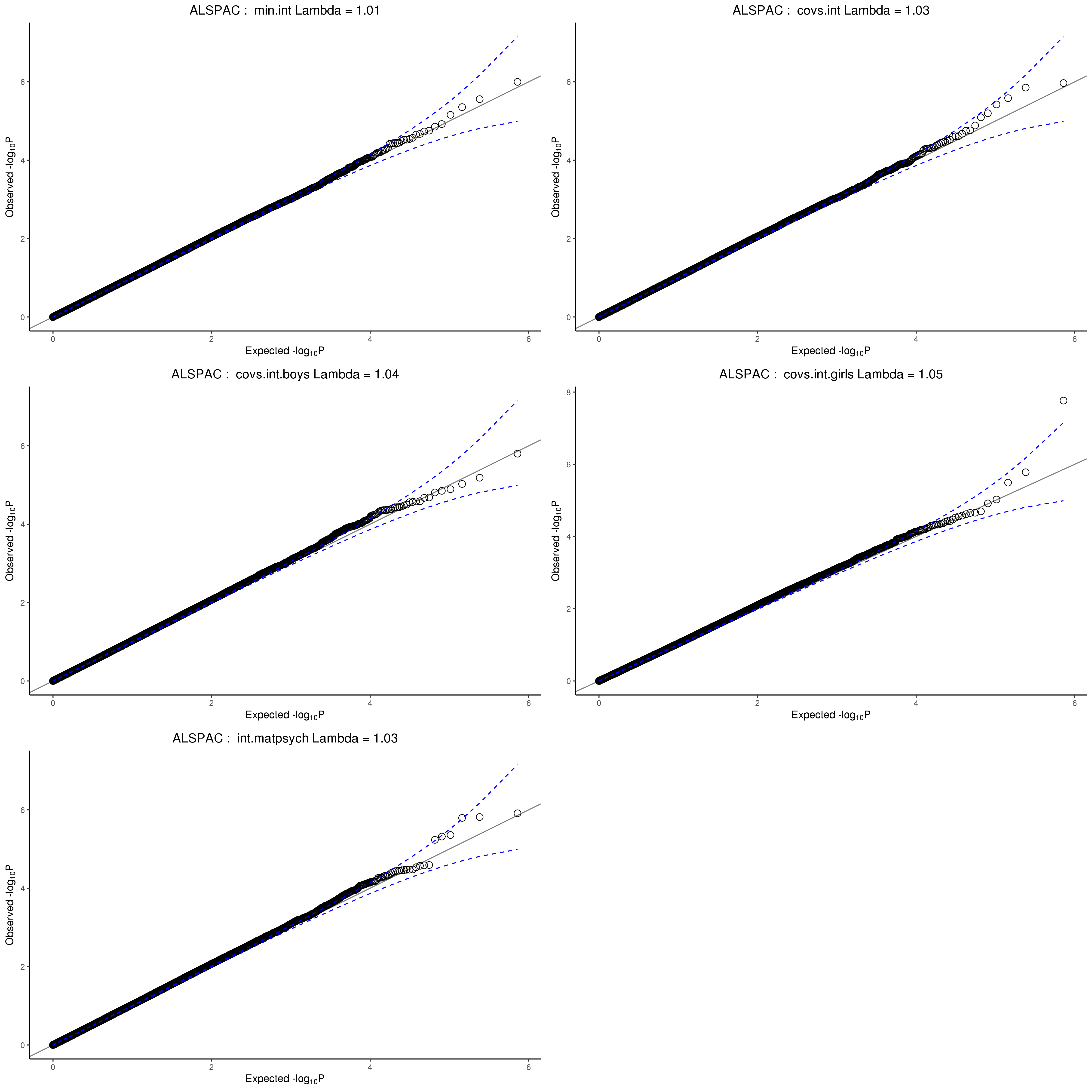
QQ-plots of the cord blood analysis age 7 – ALSPAC

Note. Quantile-Quantile (Q-Q) plot of the DNAm signals across the different models. min.int = minimally adjusted model, covs.int = covariate adjusted model, covs.int.boys = covariate, male-sex stratified model, covs.int.girls = covariate, female-sex stratified model, int.matpsych = maternal anxiety/depression adjusted model.

Figure S14
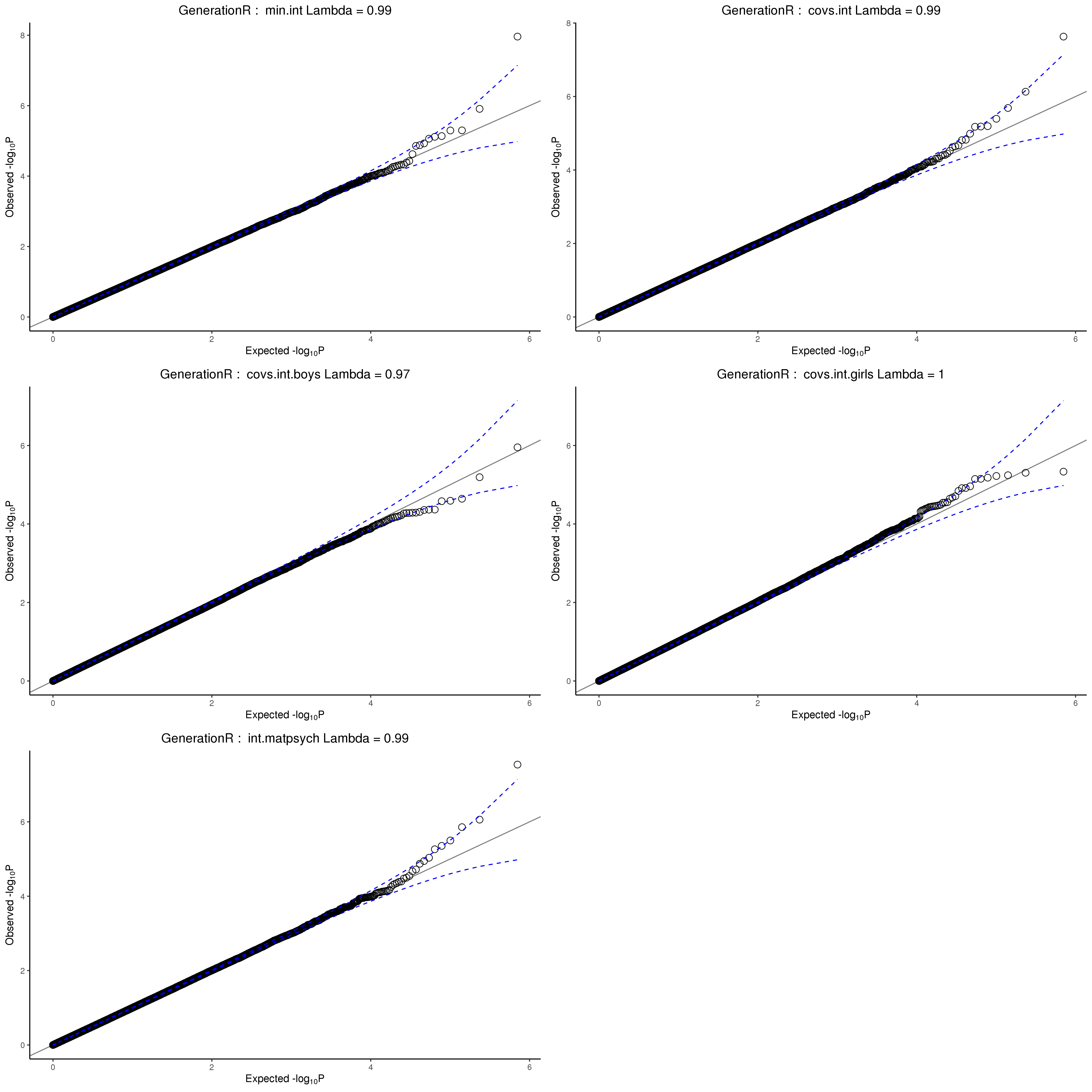
QQ-plots of the cord blood analysis age 7 – Generation R

Note. Quantile-Quantile (Q-Q) plot of the DNAm signals across the different models. min.int = minimally adjusted model, covs.int = covariate adjusted model, covs.int.boys = covariate, male-sex stratified model, covs.int.girls = covariate, female-sex stratified model, int.matpsych = maternal anxiety/depression adjusted model.

Figure S15 QQ-plots of the childhood peripheral blood analysis age 7 – ALSPAC
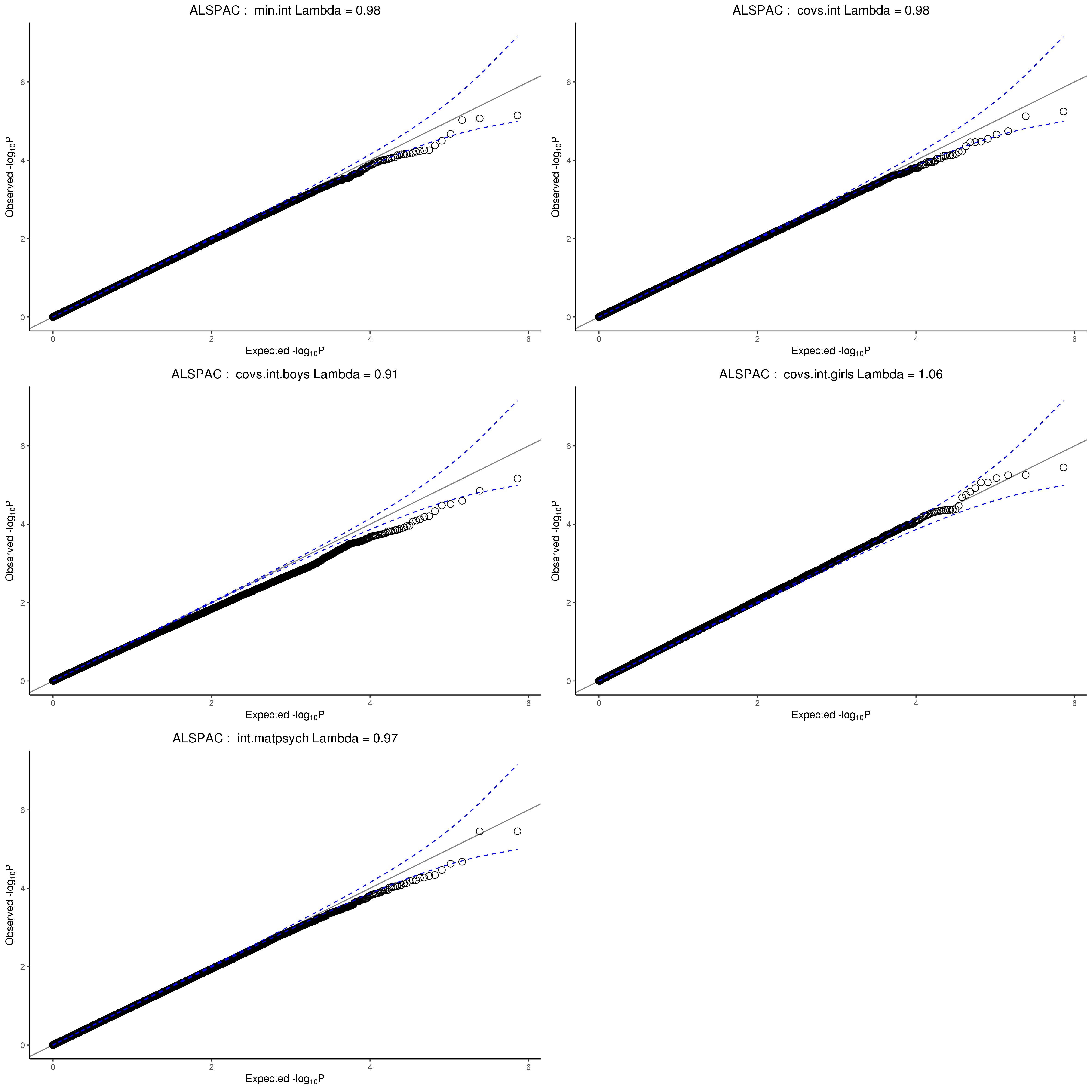

Note. Quantile-Quantile (Q-Q) plot of the DNAm signals across the different models. min.int = minimally adjusted model, covs.int = covariate adjusted model, covs.int.boys = covariate, male-sex stratified model, covs.int.girls = covariate, female-sex stratified model, int.matpsych = maternal anxiety/depression adjusted model.

Figure S16 QQ-plots of the childhood peripheral blood analysis age 7 – Generation R
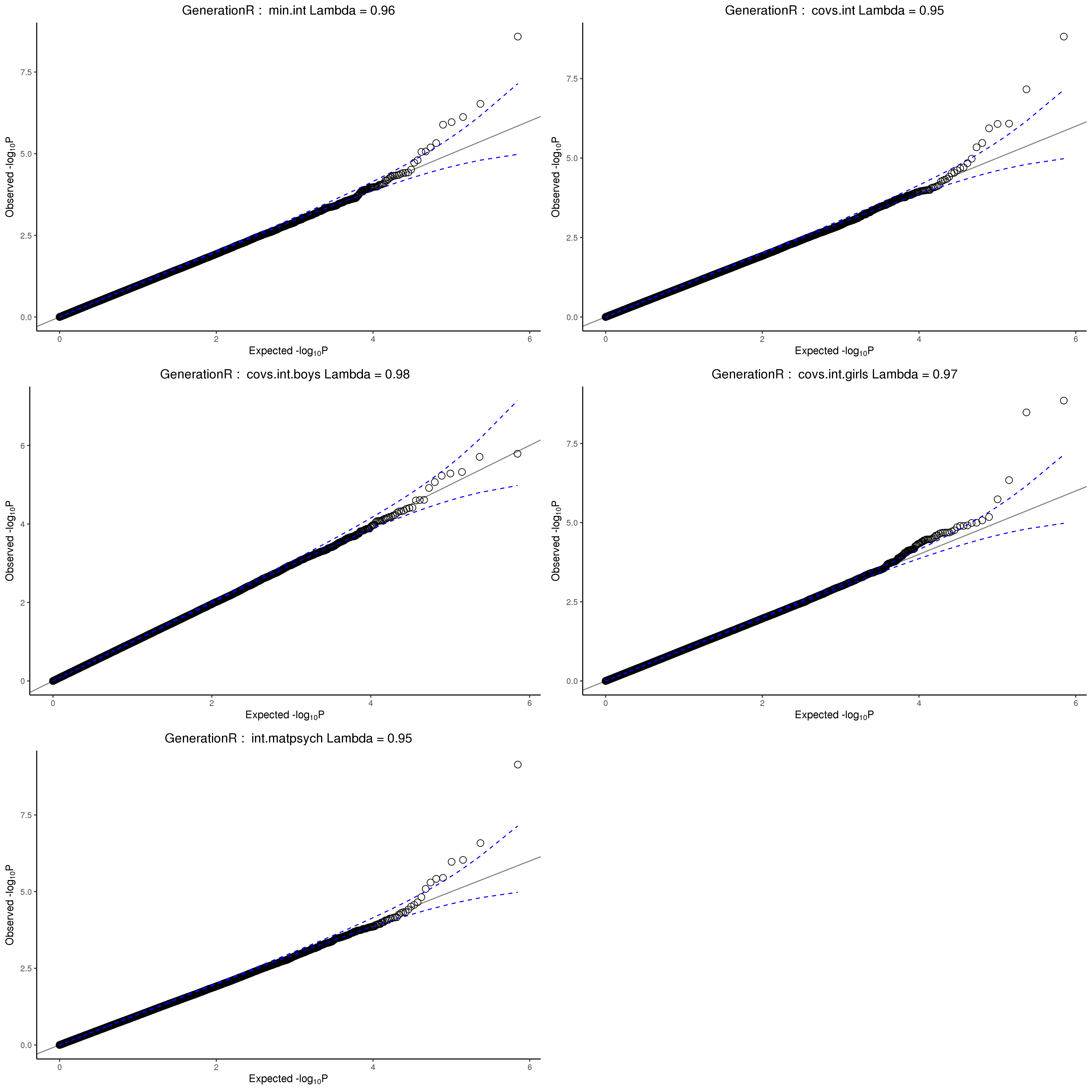

Note. Quantile-Quantile (Q-Q) plot of the DNAm signals across the different models. min.int = minimally adjusted model, covs.int = covariate adjusted model, covs.int.boys = covariate, male-sex stratified model, covs.int.girls = covariate, female-sex stratified model, int.matpsych = maternal anxiety/depression adjusted model.

##### Meta-analysis results

Correlation matrices of the models effect estimates of the meta-analysis were in line with the correlations observed in the individual cohorts (Figure S17 to S19). Overall, estimates correlated highly, except for the sex stratified model coefficients, which only correlated moderately with the other model estimates, and did not show any association with each other. This pattern was consistently observed within cohorts, across tissues, as well as in the meta-analysed results, and could indicate potential sex-differences in the association between DNA methylation and internalising problems. Alternatively, this may also be explained by residual confounding of sex (Yousefi et al., 2015) and/or the small sample size in the sex-stratified analyses. Spurious results in sex-stratified EWAS models have previously been reported in a study where offspring sex was less likely to be a confounder/important source of variation in the trait of interest (paternal BMI) (Sharp et al., 2021). Visual inspection of QQ-plots indicates that the P-values are mostly normally distributed Figures Figure S20 to S22). The plots of the leave-one-out analysis can be found in Figure S23 to Figure S25 .

Figure S17 Correlation matrix of the cord blood probe-level meta-analysis results: Internalising problems age 3
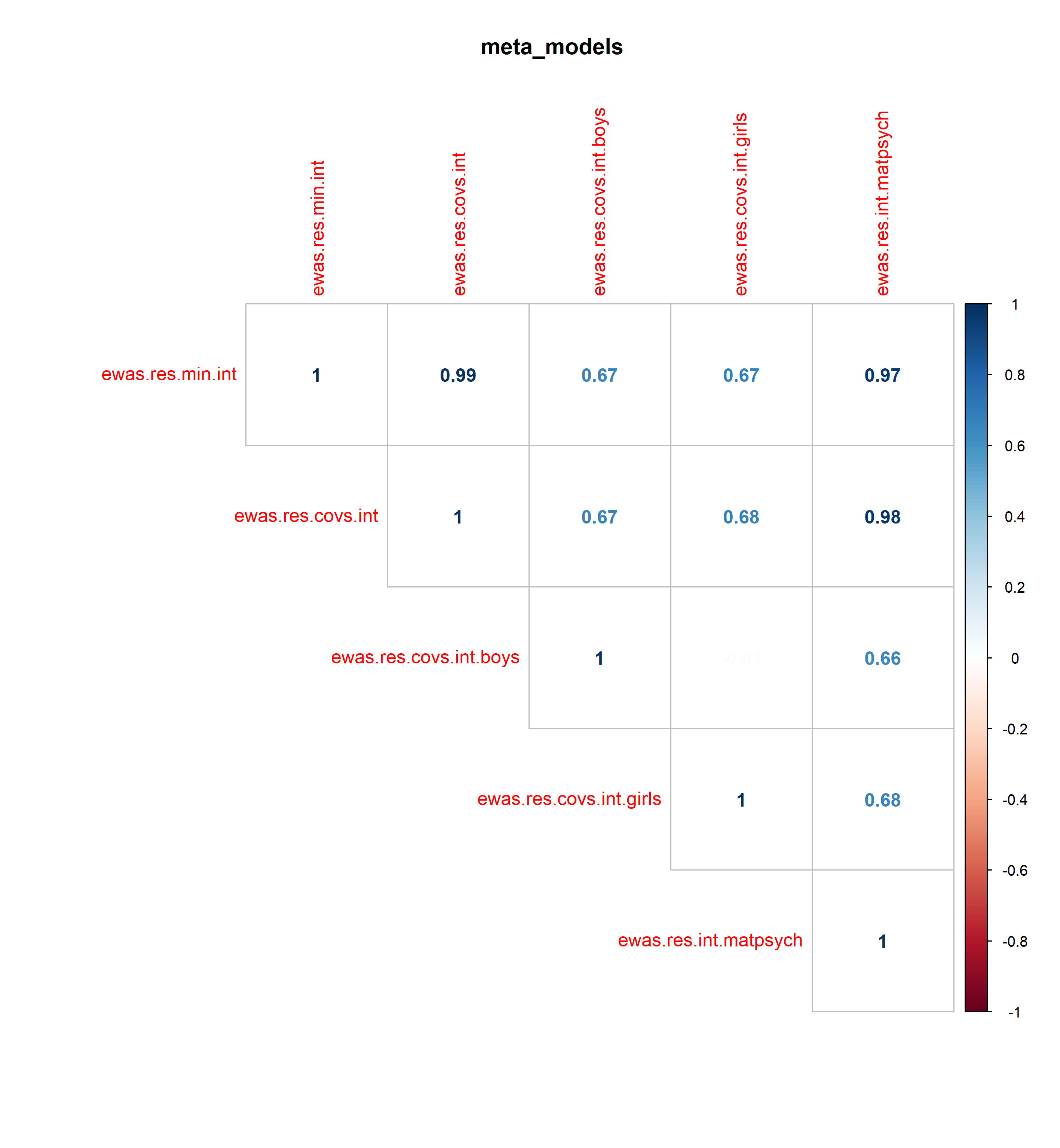

Figure S18 Correlation matrix of the cord blood probe-level meta-analysis results: Internalising problems age 7

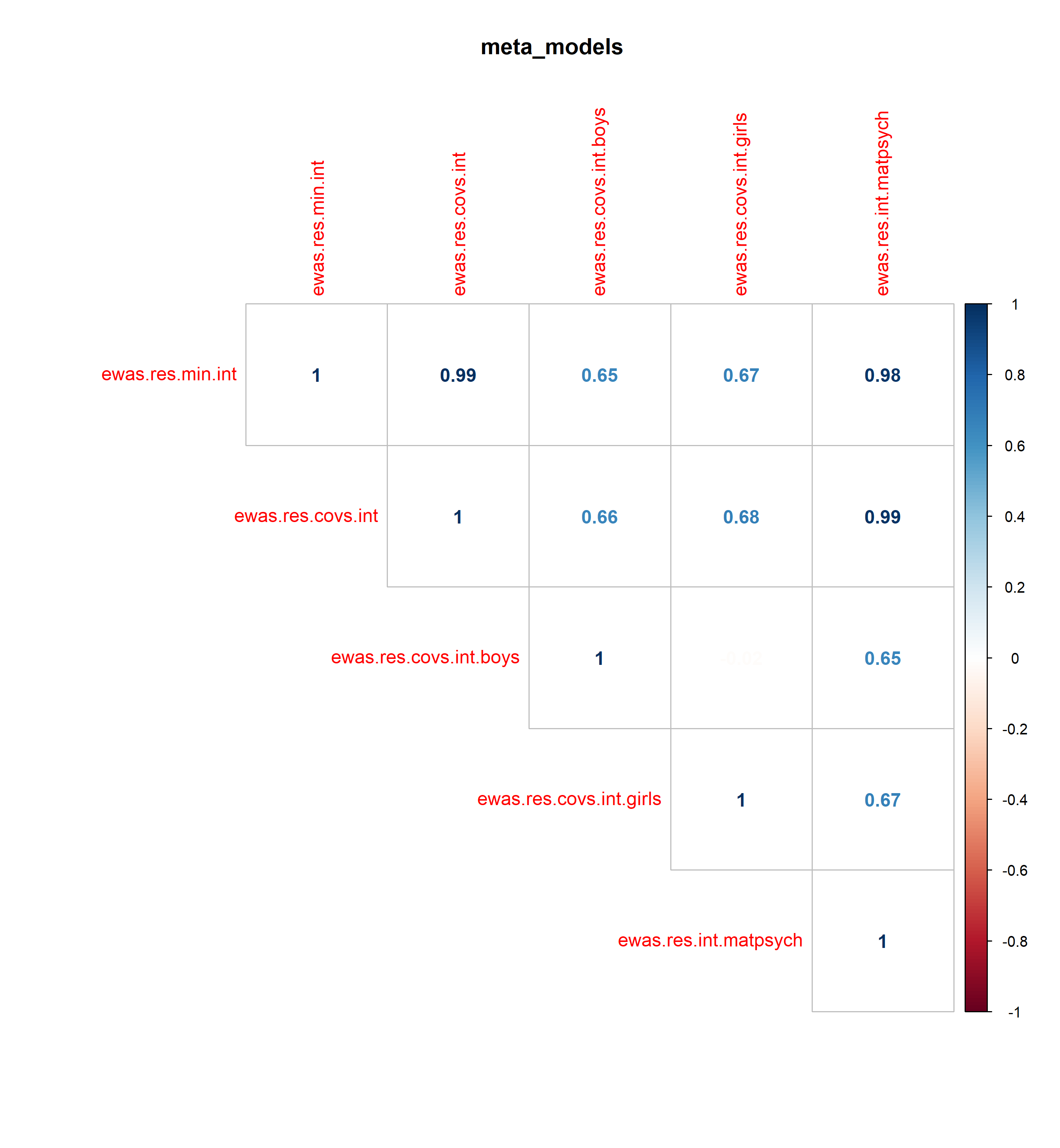

Figure S19 Correlation matrix of the cross-sectional meta-analysis results: Internalising problems age 7

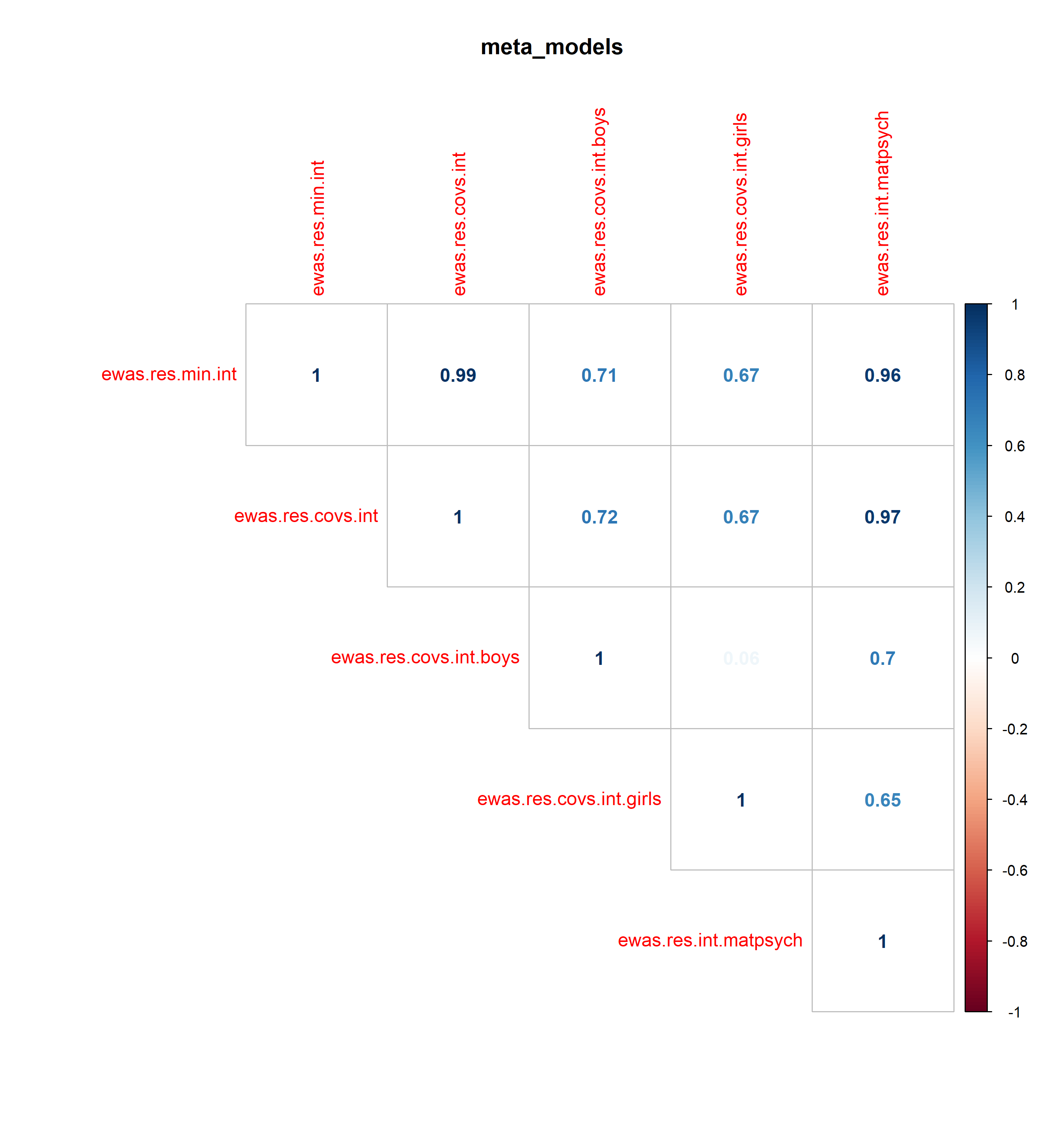

Figure S20 QQ-plot of the cord blood probe-level meta-analysis results: Internalising problems age 3

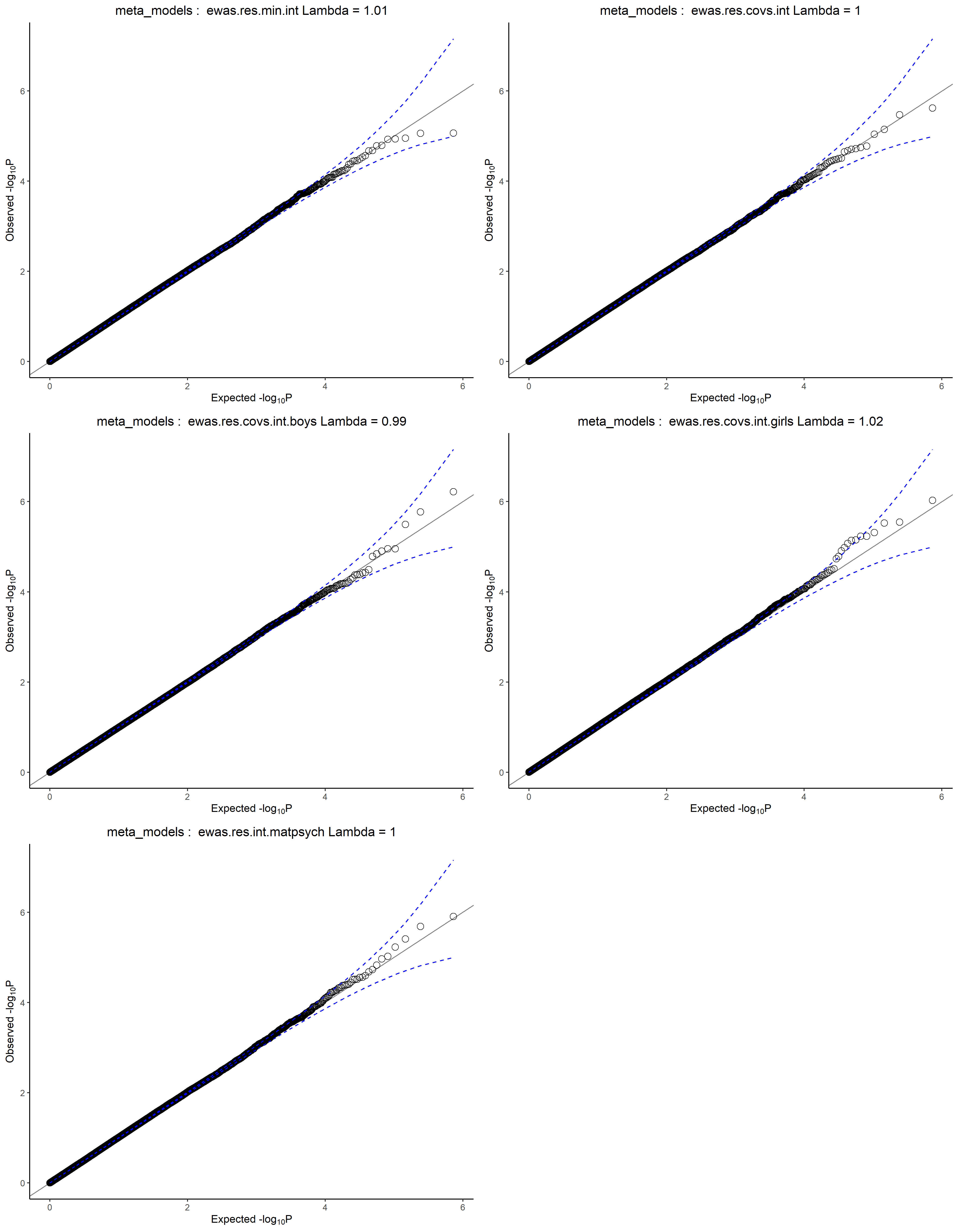

Figure S21
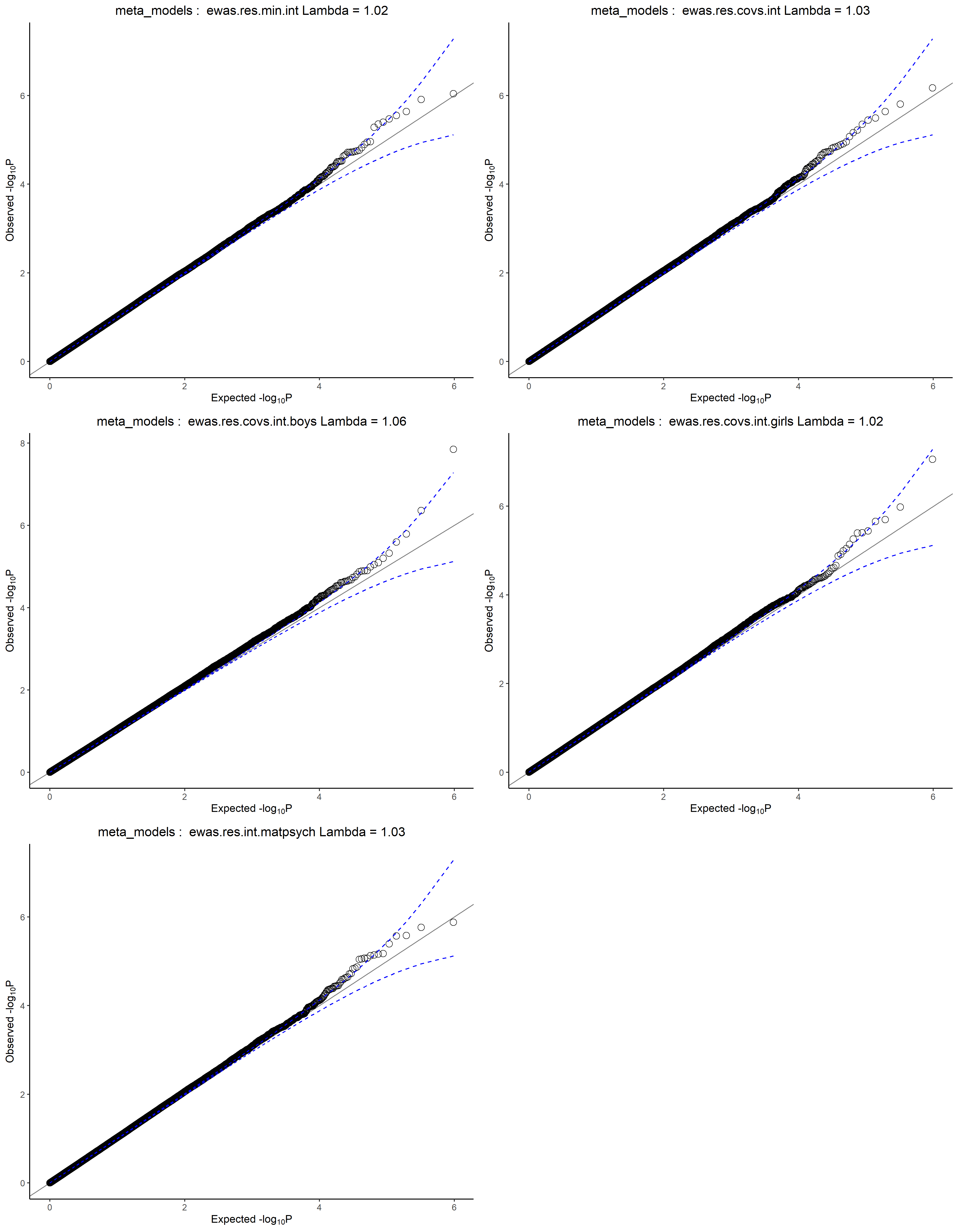
QQ-plot of the cord blood probe-level meta-analysis results: Internalising problems age 7
Figure S22 QQ-plot of the cord blood cross-sectional meta-analysis results: Internalising problems age 7

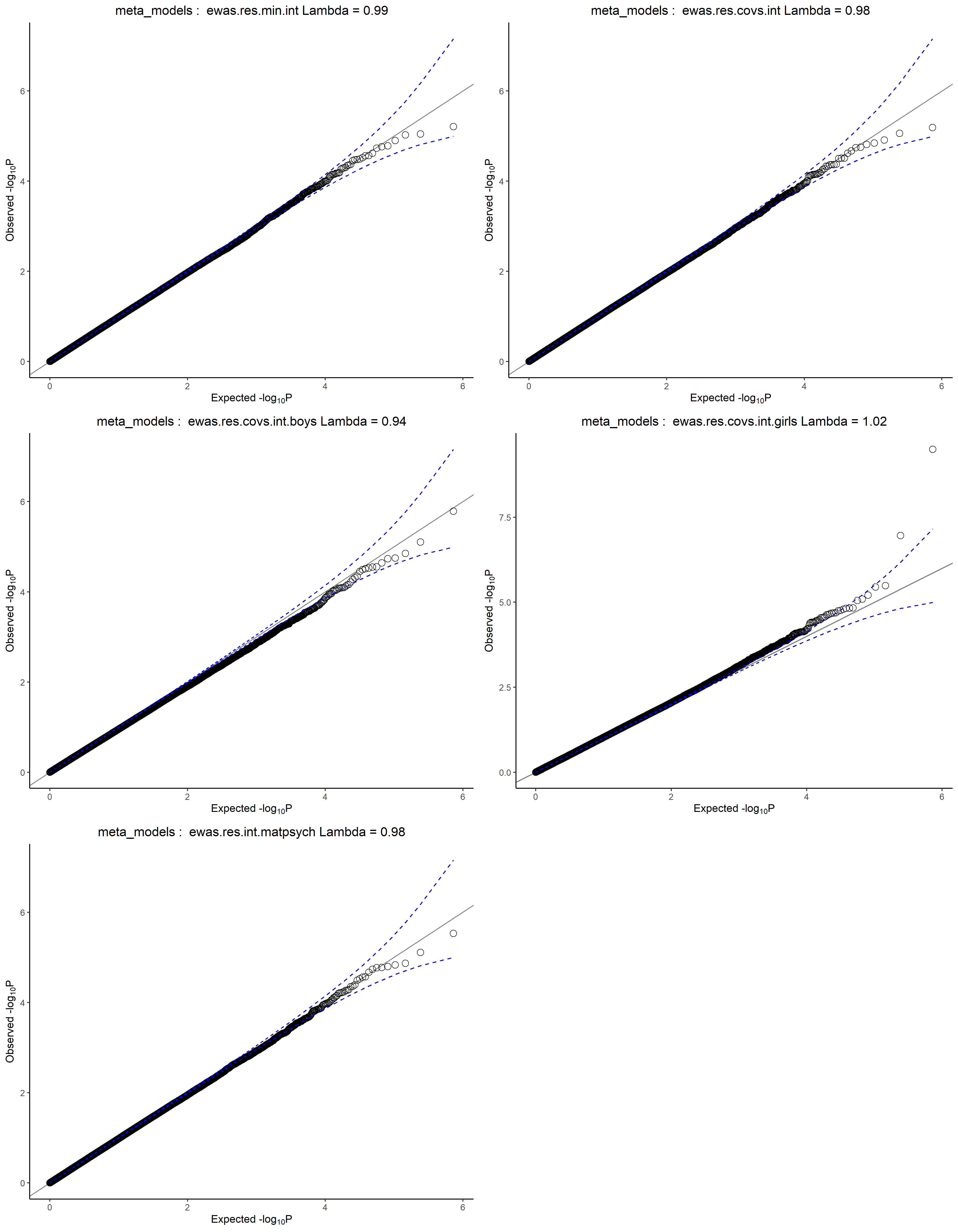

Figure S23 Leave-one-out plot of the cord-blood meta-analysis age 7	(Cg26668632)
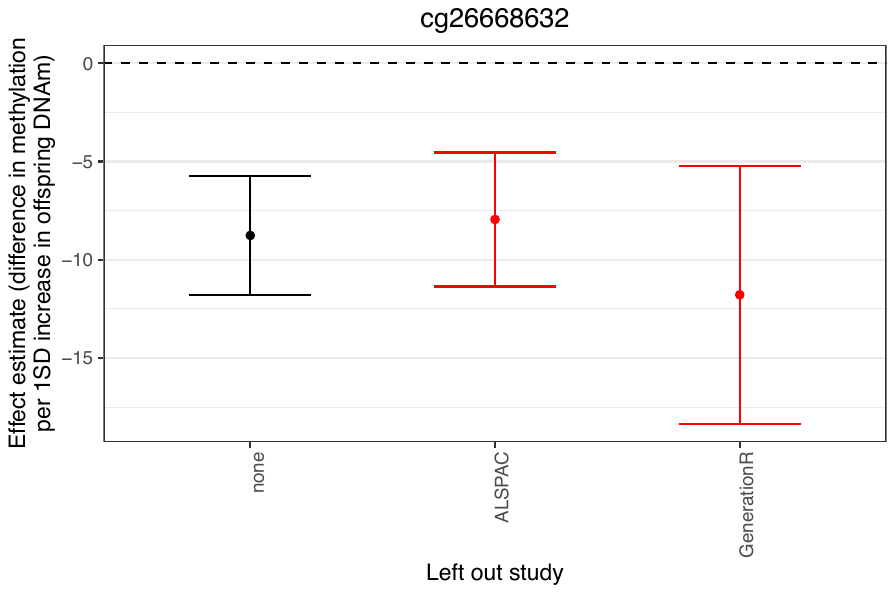

Figure S24 Leave-one-out plot of the childhood peripheral blood meta-analysis age 7 (Cg08884410)

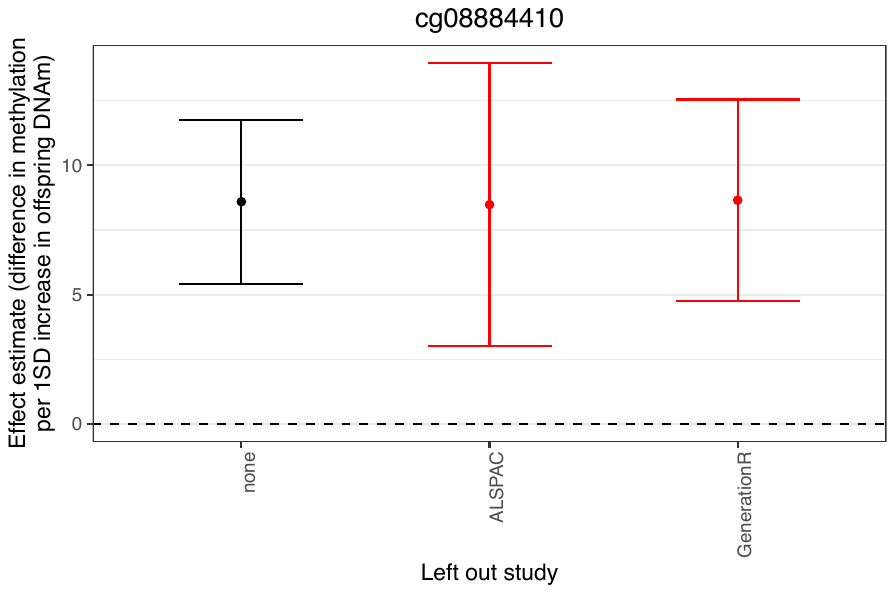

Figure S25 Leave-one-out plot of the childhood peripheral blood meta-analysis age 7 (Cg07283896)
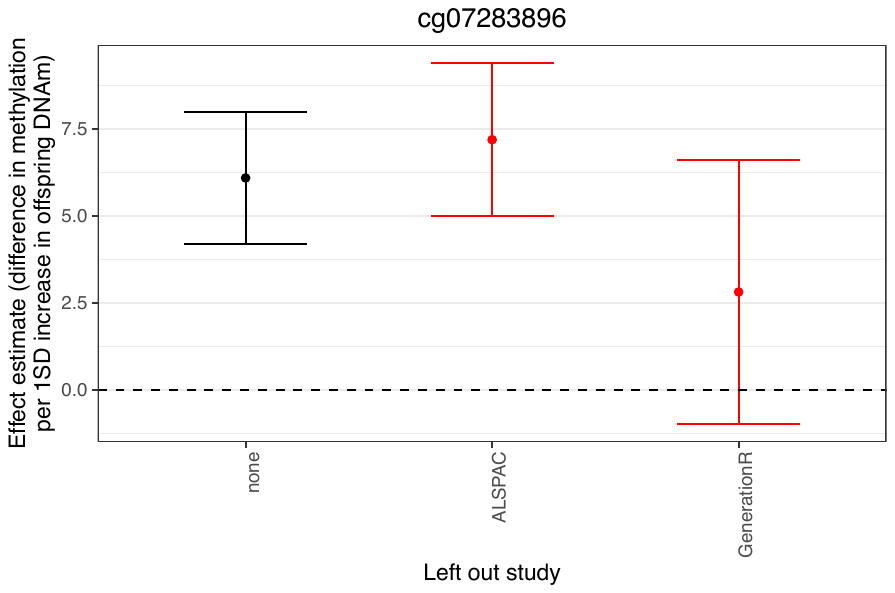

Figure S26 Manhattan and QQ-Plots of the covariate adjusted models across time-points
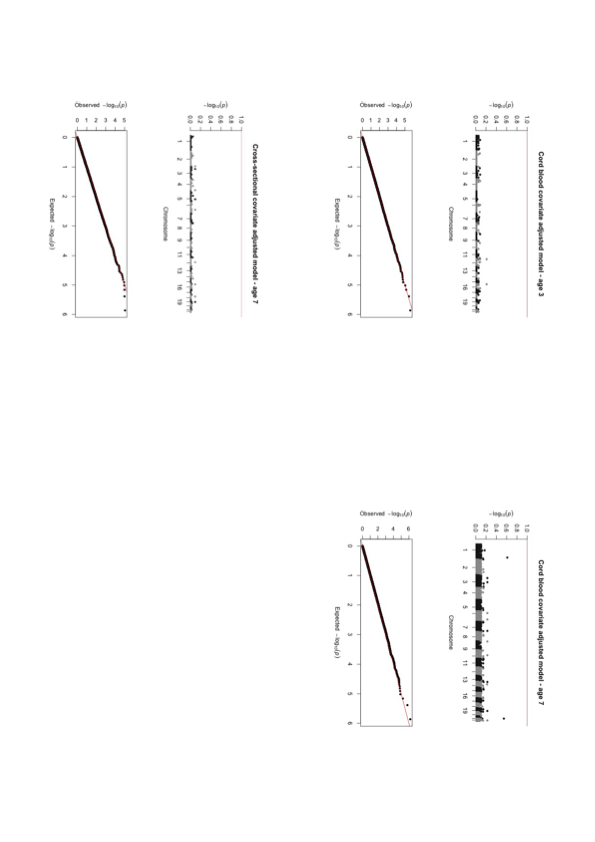
.
Figure S27
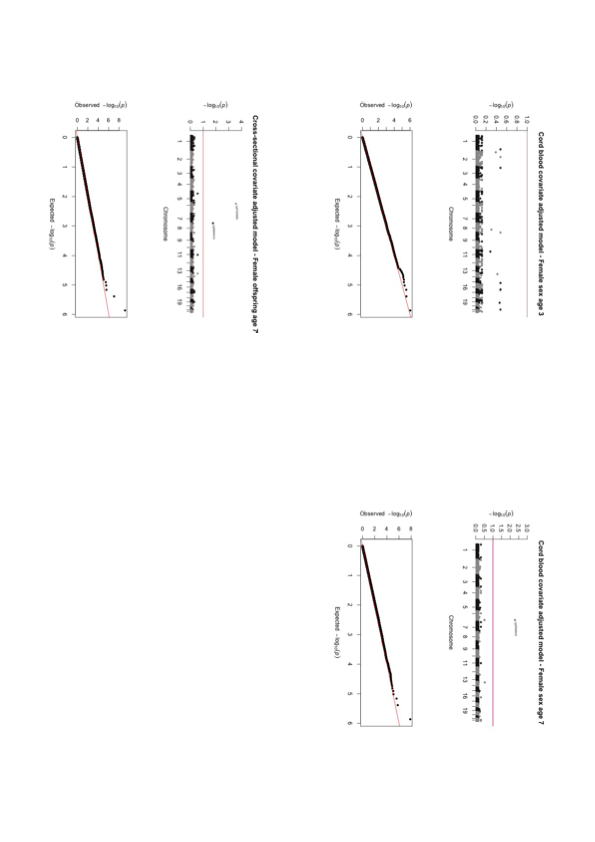
Manhattan and QQ-Plots of the covariate adjusted female-sex stratified models across time-points.
Figure S28 Manhattan and QQ-Plots of the covariate adjusted male-sex stratified models across time-points.
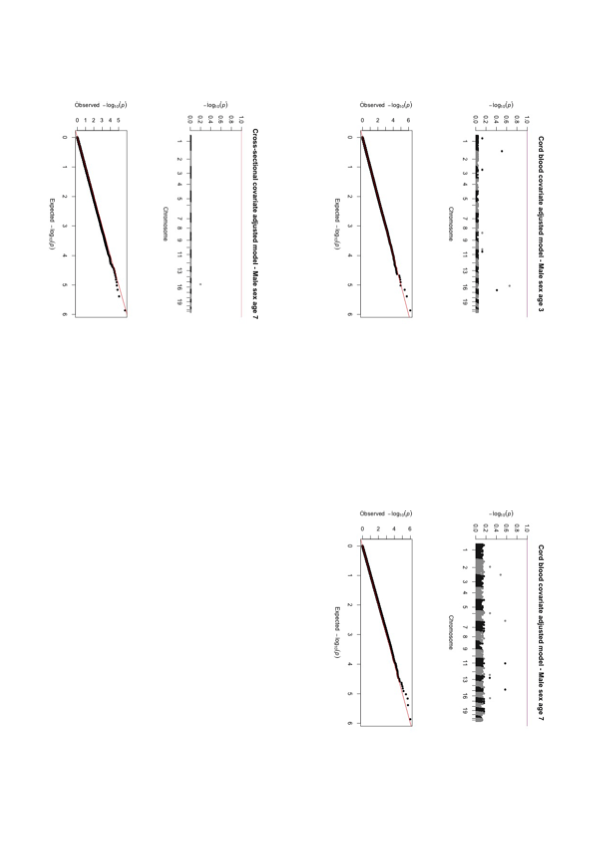

Figure S29 Manhattan and QQ-Plots of the maternal anxiety/depression adjusted models across time-points.
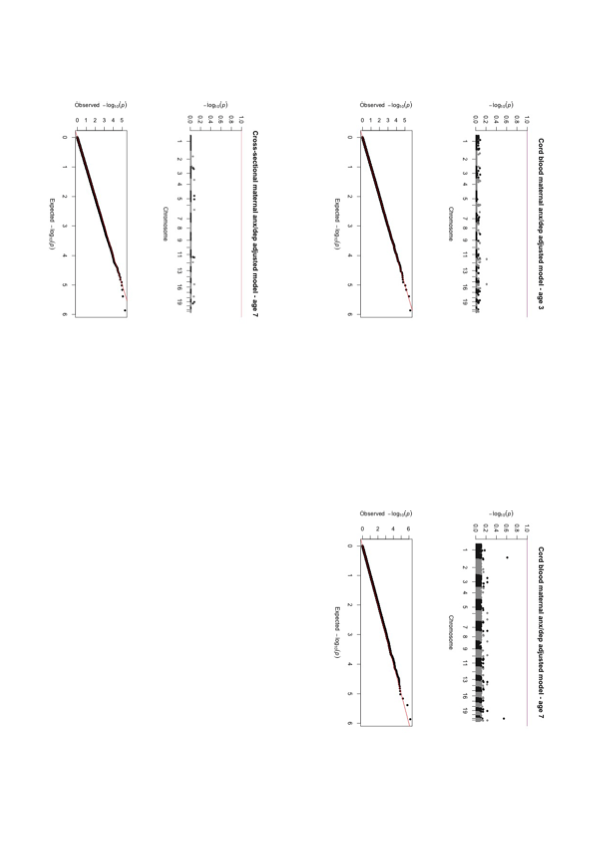

Figure S30 Correlation table of effect estimates

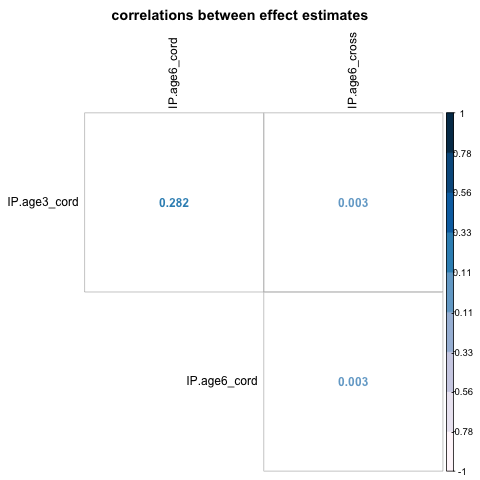

Note. IP = Internalising problems. Cord = DNAm assessed in cord blood. Cross = cross sectional analysis using DNAm assessed in peripheral blood in childhood.

Table S1 Correlation table of internalising problems and covariates

|  | Correlation with int. problems age 3 | | | | Correlation with int. problems age | |
| --- | --- | --- | --- | --- | --- | --- |
| Covariates | ALSPAC | Generation R | MoBa1 | MoBa2 | ALSPAC | Generation R |
| Maternal anxiety/depression during pregnancy | 0.13** | 0.23** | 0.19** | 0.19** | 0.15** | 0.20** |
| Maternal education | -0.03 | -0.06 | -0.01 | -0.02 | 0.05 | -0.03 |
| Maternal age | -0.05 | -0.02 | -0.07* | -0.08 | -0.01 | -0.07* |
| Gestational age | -0.02 | 0.01 | -0.06 | -0.05 | -0.03 | -0.05 |
| Internalising problems age 3 | - | - | - | - | 0.39** | 0.56** |

Note. M = mean. int problems = internalising problems. * P-value < 0.05. ** P-value < 0.001.

Table S2 Maternal smoking during pregnancy and offspring internalising problems

| Cohort | M int. problems age 3 (SD) | | M int.problems age 7 (SD) | |
| --- | --- | --- | --- | --- |
|  | No smoking or quitting early in pregnancy | Smoking late/throughout pregnancy | No smoking or quitting early in pregnancy | Smoking late/throughout pregnancy |
| ALSPAC | 0.25 (0.27) | 0.26 (0.29) | 0.25 (0.28) | 0.26 (0.29) |
| Generation R | 0.12  (0.11) | 0.12  (0.10) | 0.14  (0.14) | 0.14  (0.12) |
| MoBa1 | 0.23 (0.22) | 0.25 (0.19) | - | - |
| MoBa2 | 0.23 (0.22) | 0.29 (0.23) | - | - |

Note. M = mean. int problems = internalising problems.

Table S3 Probe-level EWAS meta-analysis results across sex and time

|  |  |  |  | **Prospective analyses (cord blood DNAm)** | | | | | | | | **Cross-sectional analysis (peripheral blood DNAm; )** | | | |
| --- | --- | --- | --- | --- | --- | --- | --- | --- | --- | --- | --- | --- | --- | --- | --- |
|  |  |  |  | **Internalising problems age 3 –**  **female sex** | | | | **Internalising problems age 6 –**  **female sex (n = 804)** | | | | **Internalising problems age 6 –**  **female sex (n = 572)** | | | |
| **CpG** | **Gene** | **Chr** | **Position** | ***n*_cohorts_ (direction)** | ***B* (*SE*)** | ***p*** | ***I^2^*** | **n_cohorts_ (direction)** | ***B* (*SE*)** | ***p*** | ***I^2^*** | **n_cohorts_ (direction)** | ***B* (*SE*)** | ***p*** | ***I^2^*** |
| cg26668632 | *IFNGR1* | Chr6 | 137540814 | 4 (--+-) | -3.97 (1.69) | 0.02 | 67.6 | 2 (--) | -8.76 (1.54) | 1.42 x 10-^08^ | 0 | 2 (--) | -4.83 (2.32) | 0.037 | 0 |
| cg07283896 | - | Chr6 | 11855283 | 4(--++) | -0.38 (0.46) | 0.41 | 30.0 | 2 (++) | 0.61 (0.47) | 0.19 | 0 | 2 (++) | 6.10 (0.97) | 3.20x 10^-10^ | 73.90 |
| cg08884410 | - | chr7 | 152620090 | 4 (++++) | 1.37 (1.06) | 0.20 | 0 | 2 (++) | 0.42 (1.06) | 0.69 | 0 | 2 (++) | 8.59 (1.62) | 1.11 x 10^-07^ | 0 |
|  |  |  |  | **Internalising problems age 3 –**  **male sex** | | | | **Internalising problems age 6 –**  **male sex (n = 797)** | | | | **Internalising problems age 6 –**  **male sex (n = 564)** | | | |
| cg26668632 | *IFNGR1* | Chr6 | 137540814 | --++ | -1.81 (1.68) | 0.28 | 49.2 | ++ | 3.71 (1.86) | 0.05 | 3 | ++ | 3.71 (1.86) | 0.05 | 3 |
| cg07283896 | - | Chr6 | 11855283 | +-+- | 0.22 (0.44) | 0.61 | 31.8 | 2 (++) | 6.10 (0.97) | 3.20x 10^-10^ | 73.90 | -+ | -0.25 (1.26) | 0.84 | 66.2 |
| cg08884410 | - | chr7 | 152620090 | +---- | -0.55 (1,01) | 0.59 | 0 | 2 (++) | 8.59 (1.62) | 1.11 x 10^-07^ | 0 | ++ | 1.26 (1.66) | 0.45 | 0 |

*Note. Cohorts = ALSPAC, Generation R, Moba 1, Moba 2. SE = Standard Error, Chr = Chromosome*

### Cohort-specific acknowledgments & funding

#### ALSPAC

We are extremely grateful to all the families who took part in this study, the midwives for their help in recruiting them, and the whole ALSPAC team, which includes interviewers, computer and laboratory technicians, clerical workers, research scientists, volunteers, managers, receptionists and nurses. The UK Medical Research Council and Wellcome (Grant ref: 217065/Z/19/Z) and the University of Bristol provide core support for ALSPAC. This publication is the work of the authors and Laura Schellhas will serve as guarantors for the contents of this paper. A comprehensive list of grants funding is available on the ALSPAC website (http://www.bristol.ac.uk/alspac/external/documents/grant-acknowledgements.pdf).

This research was performed in the UK Medical Research Council Integrative Epidemiology Unit (grant number: MC_UU_00011/7 and MC_UU_00011/5) and also supported by the National Institute for Health Research (NIHR) Bristol Biomedical Research Centre at University Hospitals Bristol NHS Foundation Trust and the University of Bristol.

#### Generation R

The Generation R Study is conducted by Erasmus MC, University Medical Center Rotterdam, in close collaboration with the School of Law and Faculty of Social Sciences of the Erasmus University Rotterdam, the Municipal Health Service Rotterdam area, Rotterdam, the Rotterdam Homecare Foundation, Rotterdam and the Stichting Trombosedienst & Artsenlaboratorium Rijnmond (STAR-MDC), Rotterdam. We gratefully acknowledge the contribution of children and parents, general practitioners, hospitals, midwives and pharmacies in Rotterdam. The study protocol was approved by the Medical Ethical Committee of the Erasmus Medical Centre, Rotterdam. Written informed consent was obtained for all participants. The generation and management of the Illumina 450K methylation array data (EWAS data) for the Generation R Study was executed by the Human Genotyping Facility of the Genetic Laboratory of the Department of Internal Medicine, Erasmus MC, the Netherlands. We thank Mr. Michael Verbiest, Ms. Mila Jhamai, Ms. Sarah Higgins, Mr. Marijn Verkerk and Dr. Lisette Stolk for their help in creating the EWAS database. We thank Dr. A.Teumer for his work on the quality control and normalization scripts.

The general design of the Generation R Study is made possible by financial support from the Erasmus MC, Erasmus University Rotterdam, the Netherlands Organization for Health Research and Development and the Ministry of Health, Welfare and Sport. The EWAS data were funded by a grant to VWJ from the Netherlands Genomics Initiative (NGI)/Netherlands Organisation for Scientific Research (NWO) Netherlands Consortium for Healthy Aging (NCHA; project nr. 050-060-810), by funds from the Genetic Laboratory of the Department of Internal Medicine, Erasmus MC, and by a grant from the National Institute of Child and Human Development (R01HD068437). V.W.J. received a Consolidator Grant from the European Research Council (ERC-2014-CoG-648916). This project received funding from the European Union’s Horizon 2020 research and innovation programme (733206, LifeCycle; 874739, LongITools; 874583, ATHLETE; 824989, EUCAN-Connect) and from the European Joint Programming Initiative “A Healthy Diet for a Healthy Life” (JPI HDHL, NutriPROGRAM project, ZonMw the Netherlands no.529051022 and PREcisE project ZonMw the Netherlands no.529051023).

#### MoBa

We are grateful to all the participating families in Norway who take part in this on-going cohort study.

We thank the Norwegian Institute of Public Health (NIPH) for generating high-quality genomic data. This research is part of the HARVEST collaboration, supported by the Research Council of Norway (#229624). We also thank the NORMENT Centre for providing genotype data, funded by the Research Council of Norway (#223273), South East Norway Health Authorities and Stiftelsen Kristian Gerhard Jebsen. We further thank the Center for Diabetes Research, the University of Bergen for providing genotype data and performing quality control and imputation of the data funded by the ERC AdG project SELECTionPREDISPOSED, Stiftelsen Kristian Gerhard Jebsen, Trond Mohn Foundation, the Research Council of Norway, the Novo Nordisk Foundation, the University of Bergen, and the Western Norway Health Authorities.

The Norwegian Mother, Father and Child Cohort Study is supported by the Norwegian Ministry of Health and Care Services and the Ministry of Education and Research. For this work, MoBa 1 and 2 were supported by the Intramural Research Program of the NIH, National Institute of Environmental Health Sciences (Z01-ES-49019) and the Norwegian Research Council/BIOBANK (grant no 221097). This work was partly supported by the Research Council of Norway through its Centres of Excellence funding scheme, project number 262700. Where authors are identified as personnel of the International Agency for Research on Cancer / World Health Organization, the authors alone are responsible for the views expressed in this article and they do not necessarily represent the decisions, policy or views of the International Agency for Research on Cancer / World Health Organization.

### Author-specific funding statements

LS: This research was also conducted as part of the CAPICE (Childhood and Adolescence Psychopathology: unravelling the complex etiology by a large Interdisciplinary Collaboration in Europe) project, funded by the European Union’s Horizon 2020 research and innovation programme, Marie Sklodowska Curie Actions – MSCA-ITN-2016 – Innovative Training Networks under grant agreement number 721567. This study was supported by the NIHR Biomedical Research Centre at the University Hospitals Bristol NHS Foundation Trust and the University of Bristol. The views expressed in this publication are those of the authors and not necessarily those of the NHS, the

National Institute for Health Research or the Department of Health and Social Care.

GCS is financially supported by an MRC New Investigator Research (grant code MR/S009310/1), an MRC project grant (MR/W020297/1) and the European Joint Programming Initiative “A Healthy Diet for a Healthy Life” (JPI HDHL, NutriPROGRAM project, UK MRC MR/S036520/1)

LZ is financially supported by the Health Data Science Centre, Fondazione Human Technopole, Milan, Italy, and by the Data and Connectivity National Core Study, led by Health Data Research UK in partnership with the Office for National Statistics and funded by UK Research and Innovation (grant ref MC_PC_20058), with additional support by The Alan Turing Institute via ‘Towards Turing 2.0’ EPSRC Grant Funding.

CAMC is supported by the European Union's Horizon 2020 Research and Innovation Programme (EarlyCause, grant agreement No 848158), the HorizonEurope Research and Innovation Programme (FAMILY, grant agreement No 101057529; HappyMums, grant agreement No 101057390) and the European Research Council (TEMPO; grant agreement No 101039672). This research was conducted while CAMC was a Hevolution/AFAR New Investigator Awardee in Aging Biology and Geroscience Research.

AH was supported by the HorizonEurope Research and Innovation Programme (FAMILY, grant agreement No 101057529) and the South-Eastern Norway Regional Health Authority (#2020022).

MB: This work was also supported by the Research Council of Norway grant number 288083 and 301004
